# Group Testing Large Populations for SARS-CoV-2

**DOI:** 10.1101/2021.06.03.21258258

**Authors:** Hooman Zabeti, Nick Dexter, Ivan Lau, Leonhardt Unruh, Ben Adcock, Leonid Chindelevitch

## Abstract

Group testing, the testing paradigm which combines multiple samples within a single test, was introduced in 1943 by Robert Dorfman. Since its original proposal for syphilis screening, group testing has been applied in domains such as fault identification in electrical and computer networks, machine learning, data mining, and cryptography. The SARS-CoV-2 pandemic has led to proposals for using group testing in its original context of identifying infected individuals in a population with few tests. Studies suggest that non-adaptive group testing - in which all the tests are determined in advance - for SARS-CoV-2 could help save 20% to 90% of tests depending on the prevalence. However, no systematic approach for comparing different non-adaptive group testing strategies currently exists.

In this paper we develop a software platform for evaluating non-adaptive group testing strategies in both a noiseless setting and in the presence of realistic noise sources, modelled on published experimental observations, which makes them applicable to polymerase chain reaction (PCR) tests, the dominant type of tests for SARS-CoV-2. This modular platform can be used with a variety of group testing designs and decoding algorithms. We use it to evaluate the performance of near-doubly-regular designs and a decoding algorithm based on an integer linear programming formulation, both of which are known to be optimal in some regimes. We find savings between 40% and 91% of tests for prevalences up to 10% when a small error (below 5%) is allowed. We also find that the performance degrades gracefully with noise. We expect our modular, user-friendly, publicly available platform to facilitate empirical research into non-adaptive group testing for SARS-CoV-2.

## 1. Introduction

The current COVID-19 pandemic has required a widespread mobilization of resources aimed at controlling the morbidity and mortality due to its etiological agent, SARS-CoV-2. Since the primary human-to-human transmission of SARS-CoV-2 is through respiratory droplets and contact, testing has played a key role in control efforts by enabling rapid self-isolation, contact tracing and monitoring, and limiting exposure at the population level. However, the requirements of regularly testing populations at scale have proven a challenge for many governments and public health agencies, especially in low-resource settings.

Group testing is an approach to testing introduced by Robert Dorfman in 1943 (Dorfman 1943) to provide a more efficient method of screening large numbers of men for syphilis in the United States during conscription for World War II. Since syphilis was a rare disease, most tests would produce a negative result. To improve efficiency, Dorfman proposed pooling the samples (i.e. combining them inside the same test tube) to test multiple individuals of a group at once for infection. If the result of a group test was negative, the individuals were assumed to be free of infection. If the result was positive, it was assumed that at least one individual had the infection and the members of the group would be re-tested individually to identify the infected individuals. By pooling and testing in this manner, the total number of tests to be conducted could be reduced significantly over the amount required for individual testing provided that the prevalence was low.

Testing in this manner is called *adaptive* group testing. Here, multiple rounds of testing are performed and the way samples are grouped together in any round depends on the previous ones (e.g. in Dorfman’s approach the decision about which individuals are re-tested depends on which groups were positive). In contrast, in *non-adaptive* group testing all groups are decided in advance. In testing for SARS-CoV-2, adaptive testing has two main drawbacks. First, laboratory testing of samples for the presence of viral RNA is time-consuming, taking around 4-5 hours for a single round (Ben-Ami, Klochendler, Seidel, Sido, Gurel-Gurevich, Yassour, Meshorer, Benedek, Fogel, Oiknine-Djian *et al*. 2020). However, results need to be available quickly to enable isolation of infected individuals. Non-adaptive testing takes only as long as individual testing plus the extra time required to pool samples. Second, basing successive rounds on the results of previous ones makes the scheme sensitive to human error, as results need to be repeatedly read out and interpreted. On the other hand, by deciding in advance which samples are pooled together, a non-adaptive testing procedure can be fully automated and all the tests carried out in parallel (Shental, Levy, Wuvshet, Skorniakov, Shalem, Ottolenghi, Greenshpan, Steinberg, Edri, Gillis *et al*. 2020).

For these reasons, much research has focused on *non-adaptive* group testing. However, despite many existing publications with a variety of group testing schemes, no software platform currently exists for a rigorous comparison of different approaches in a systematic way. This leaves open the question of which specific non-adaptive group testing approach provides the best balance of accuracy, cost and complexity in a particular context.

In this paper, we propose an open-source software platform, named **GroupTesting**, for evaluating and optimizing non-adaptive group testing approaches for COVID-19 testing. Our platform is robust, flexible, and modular by design. We focus on non-adaptive group testing and account for the constraints on group sizes and divisibility imposed by practical COVID-19 testing, and include realistic, tunable noise models to reflect the performance of existing COVID-19 tests. For definiteness, we use a near-doubly-regular design (also known as a constant-items-per-test and constant-tests-per-item design), but our platform can accommodate other designs seamlessly. To ensure that the determination of the infected individuals is as accurate as possible, we use an Integer Linear Programming-based method, known to be optimal in the absence of noise; however, thanks to its modular design, our platform also allows other methods to be employed by the user.

For a given user-specified set of characteristics (test performance, population size, number of tests, and estimated prevalence), our platform provides the optimal group size and the expected performance of a group testing scheme. It is available at https://github.com/WGS-TB/GroupTesting and can be installed via The Python Package Index (Pypi). To demonstrate its efficacy, we also perform a series of numerical experiments. These experiments examine the performance of group testing across a range of experimental conditions, including different prevalences (average infection rates), group sizes, and noise models.

The outline of the remainder of this paper is as follows. We commence in Section 2 with preliminary material. This includes key definitions, an overview of the relevant literature, a description of the specific measurement model and integer linear program used, as well as the group testing scheme considered. In Section 3 we discuss noise models for SARS-CoV-2 testing. Then in Section 4 we describe the software package **GroupTesting**. In Section 5 we present numerical experiments using this package, and then we conclude in Section 6 with a summary and discussion of future directions.

## 2. Preliminaries

### 2.1. Definitions

We begin by defining the key terminology used throughout the paper. These definitions are mostly in agreement with a recently published review paper (Verdun, Fuchs, Harar, Elbrächter, Fischer, Berner, Grohs, Theis, and Krahmer 2020), with appropriate notes when there are differences.

- A **group testing scheme** is a procedure for testing a population of possibly infected people for the presence or absence of an infection by using **group tests** (also known as **pool tests**). Each test combines biological samples from a subset of individuals in the population, and returns a positive result if and only if a least one of them is infected.
- The **population size** *N* is the number of people we wish to test. We typically assume 100 ≤ *N* ≤ 10, 000.
- The **number of tests** *m* is the overall number of tests (pools, groups) used in a group testing scheme. We try to minimize this number to improve the efficiency of the scheme.
- In an **adaptive** group testing scheme, some of the testing decisions (i.e. which members of the population combine into each group test) are taken after learning the results of a subset of the tests, while in a **non-adaptive** group testing scheme, all the testing decisions are made in advance. We focus on non-adaptive group testing schemes here.
- A non-adaptive group testing scheme is comprised of two parts. First, a **design matrix** (also called **test design**) is the binary matrix ***A*** ∈ {0, 1}^*m×N*^ with a row for each test and a column for each member of the population, such that ***A***_*ij*_ = 1 if and only if test *i* includes individual *j*. Second, a **decoding algorithm** specifies how the **outcome vector *y*** ∈ {0, 1}^*m*^, with *y*_*i*_ = 1 if and only if test *i* is positive, is converted to a **status vector *x*** ∈ {0, 1}^*N*^, with *x*_*j*_ = 1 if and only if individual *j* is inferred to be infected.
- The **prevalence** *p* is the probability that a randomly chosen member of the population of interest is infected. We assume that *p* ≤20% in the context of COVID-19, in line with reported population-level figures (Roser, Ritchie, Ortiz-Ospina, and Hasell 2020).
- The **divisibility** *d* is the maximum number of times a person’s sample can be included in a test. We assume that *d* ≤16, in line with the fact that aliquots as small as 50 *µ*L are sufficient for pooling (Abdalhamid, Bilder, McCutchen, Hinrichs, Koepsell, and Iwen 2020) and a patient sample has a volume around 0.7 mL (Primerdesign 2020).
- The **group size** *g* is the maximum number of samples included in a single group. We assume that *g* ≤32, shown reasonable in previous work (Yelin, Aharony, Shaer-Tamar, Argoetti, Messer, Berenbaum, Shafran, Kuzli, Gandali, Hashimshony *et al*. 2020).
- The **sensitivity** is the probability of returning a correct positive result for an infected person. We distinguish between the **sensitivity of a single test**, denoted *s*_*e*_, which refers to a group containing a single individual, and the **sensitivity of a method**, denoted *S*_*e*_, which refers to an entire group testing scheme. We typically want to detect all infected people, and require either *S*_*e*_ = 1 or *S*_*e*_ ≥ 1 *− ε* for a small tolerance *ε* > 0.
- The **specificity** is the probability of returning a correct negative result for an uninfected person. As above, we distinguish between the **specificity of a single test**, denoted *s*_*p*_, and the **specificity of a method**, denoted *S*_*p*_. We typically allow a small fraction of false positive results, and tolerate values of *S*_*p*_ ≥ 0.9 in some settings.
- The **balanced accuracy** *BA* is a composite evaluation metric for a method, defined as the arithmetic mean of its sensitivity and specificity: *BA* = (*S*_*e*_ + *S*_*p*_)*/*2.
- The **number of infecteds** *K* is the total number of infected people in the population. Note that the expected value of *K* is *pN*.
- The **efficiency** *E* is the number of tests per person for a given scheme. Note that the expected value of *m* is *EN*.

### 2.2. Previous work

Since the pioneering work of Dorfman, various mathematical models have been considered for group testing in the literature (Aldridge, Johnson, and Scarlett 2019, p. 203). Some of these models, such as non-adaptive testing and noisy testing, are motivated by real-world restrictions. Prior to (Gandikota, Grigorescu, Jaggi, and Zhou 2019), the models in the literature often unrealistically assumed that a group may contain an arbitrary number of items (unbounded *g*), and an item may participate in an arbitrary number of tests (unbounded *d*). (Gandikota *et al*. 2019) introduce the sparsity-constrained group testing, in which they consider *separately* the following models: (a) the finite-tests-per-item model (items have divisibility at most *d* ≤ *γ*); or (b) the finite-items-per-test model (groups have size at most *g* ≤ *ρ*). Furthermore, they consider the effect of noise on the test outcomes.

In (Gebhard, Hahn-Klimroth, Parczyk, Penschuck, Rolvien, Scarlett, and Tan 2020), it was shown that a pooling scheme based on a random regular test design together with the Definite Defectives (DD) algorithm (Aldridge, Baldassini, and Johnson 2014) is order-optimal for the finite-items-per-test model. Their simulations suggest that the decoding procedure succeeds for moderate population sizes. They also show the asymptotic optimality of DD algorithm when equipped with a suitably-chosen random test design for the finite-items-per-test model. Neither of these consider the impact of noise on the test outcomes.

To the best of our knowledge, no prior theoretical study of group testing models considers non-adaptive testing with both the finite-tests-per-item and finite-items-per-test constraints, and includes the effect of noise. In applications-based studies that use group testing to reduce the number of tests required to identify infected individuals during the COVID-19 pandemic, we argue that all of these requirements should be satisfied simultaneously.

Many studies on COVID-19 group testing use adaptive 2-stage testing (Yelin *et al*. 2020; Sinnott-Armstrong, Klein, and Hickey 2020; Abdalhamid *et al*. 2020; Hogan, Sahoo, and Pinsky 2020; Gollier and Gossner 2020; Ben-Ami *et al*. 2020; Shani-Narkiss, Gilday, Yayon, and Landau 2020; Deckert, Bärnighausen, and Kyeia 2020; Cherif, Grobe, Wang, and Kotanko 2020). Among those studies which consider a non-adaptive model (Täufer 2020; Verdun *et al*. 2020; Nalbantoglu 2020; McDermott, Stoddard, Woolf, Ellingford, Gokhale, Taylor, Demain, Newman, and Black 2021; Seong 2020; Shental *et al*. 2020), only the latter two consider noise models. However, Seong (2020) only consider a “symmetric noise” model (Aldridge *et al*. 2019, Example 3.1) defined as tests whose sensitivity *S*_*e*_ and specificity *S*_*p*_ are equal, which is unrealistic for the tests typically used for SARS-CoV-2. Shental *et al*. (2020) considered two noise models, both of which lowered test sensitivity. We summarize these studies in Table 1.

**Table 1:**
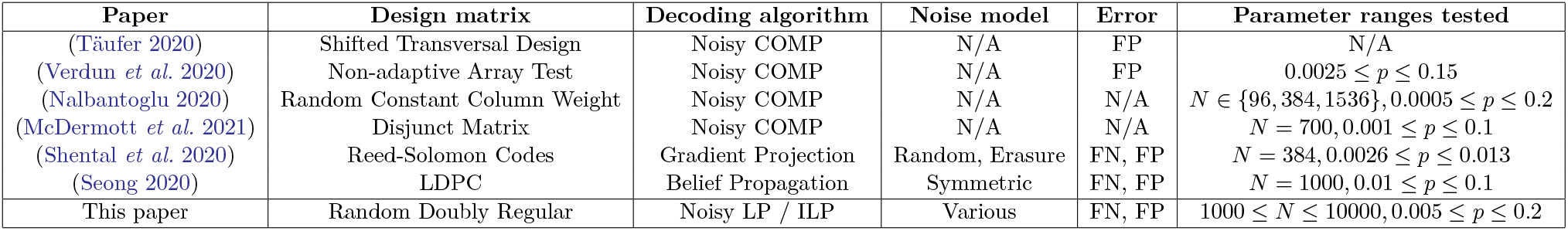
Summary of previously published non-adaptive group testing schemes for COVID-19 that satisfy the constraints we outlined. The decoding algorithms and the noise models are discussed in (Aldridge *et al*. 2019, §3). Here ‘FP’ and ‘FN’ stand for False Positive and False Negative respectively. For further discussion on the design matrices please see Section 2.4.

**Table 2:** Default values of noise model parameters in **GroupTesting**

|  |  |
| --- | --- |
| $\underline{\theta}$ | 0.0 |
| $\overline{\theta}$ | 0.0625 |
| $P_{\text{FN}}$ | 0.1 |
| $\rho$ | 0.01 |

We note that besides the decoding algorithms listed in Table 1, other decoding algorithms have been previously used in the literature, such as those in (Aldridge *et al*. 2019, §3). In addition, a recent paper (Ciampiconi, Ghosh, Scarlett, and Meel 2020) proposes a MaxSAT framework for decoding, whose current implementation uses a hybrid approach leveraging a satisfiability solver as well as an integer linear programming solver like the one we adopt here.

Finally, we also note that standard group testing, where test results are binary (positive or negative), is not the only framework proposed for efficiently identifying infected COVID-19 individuals. A closely related, but less standard group testing approach based on compressed sensing (Foucart and Rauhut 2013) has also been proposed (Ghosh, Rajwade, Krishna, Gopalkrishnan, Schaus, Chakravarthy, Varahan, Appu, Ramakrishnan, Ch *et al*. 2020b; Ghosh, Agarwal, Rehan, Pathak, Agrawal, Gupta, Consul, Gupta, Goyal, Rajwade *et al*. 2020a; Zhu, Rivera, and Baron 2020; Yi, Mudumbai, and Xu 2020c; Petersen, Bah, and Jung 2020; Yi, Cho, Wu, Mudumbai, and Xu 2020a; Yi, Cho, Wu, Xu, and Mudumbai 2020b; Cohen, Shlezinger, Solomon, Eldar, and Médard 2020). This approach assumes that the viral load is additive for different individuals in a given pool and that it can be accurately measured. The latter assumption does not seem realistic with standard COVID-19 testing technology, as evaluating the viral load requires additional, quantitative tests (Pujadas, Chaudhry, McBride, Richter, Zhao, Wajnberg, Nadkarni, Glicksberg, Houldsworth, and Cordon-Cardo 2020). Under these assumptions, however, the compressed sensing-based approach can not only identify whether an individual is infected, but also estimate their viral load. We return to the question of using viral loads within COVID-19 group testing in the Discussion section.

### 2.3. Group testing and measurement systems

As in the Section 2.1, let *N* be the number of individuals in the population to be tested and let *m* be the number of tests to be performed. Let ***x*** ∈ {0, 1} ^*N*^ be the status vector, where an entry of 1 in component *j* corresponds to infection in the *j*th individual. Let ***y*** ∈ **{**0, 1}^*m*^ be the outcome vector, where a value *y*_*t*_ = 1 implies that group *t* has at least one infected individual and a value of 0 implies the group is free of infection. In group testing, in the absence of noise the measurement vector ***y*** satisfies

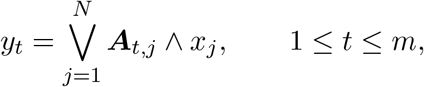

which can be more concisely represented as

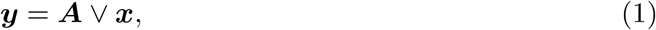

where ∨ is the Boolean inclusive `OR` operator. Let *K* be the number of infected individuals, i.e., ‖***x*** ‖_0_ = *K*, where ‖***x***‖ _0_ is the number of non-zero elements of ***x***. Assuming that the number of infected individuals is small, one can consider the Boolean vector recovery problem

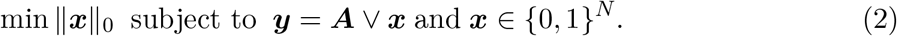

Observe that problem (2) can be restated as the following 0-1 integer linear program:

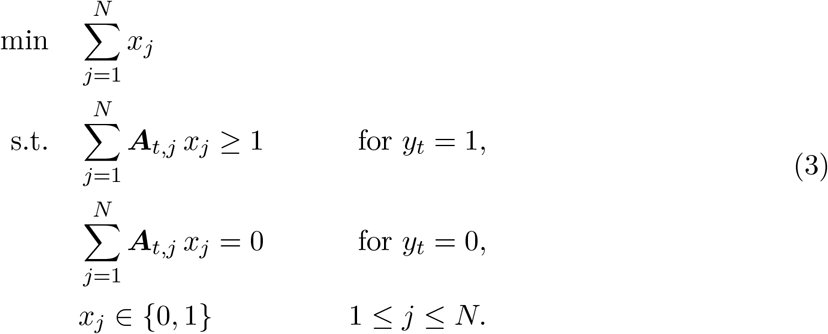

This allows us to solve the problem using integer linear program solvers such as IBM ILOG CPLEX Optimizer (**CPLEX**) (IBM 2020), **Gurobi** (Gurobi Optimization 2021), and the GNU Linear Programming Kit (**GLPK**) (Makhorin). This problem is NP-hard (Aldridge 2019, Remark 2.1); in fact, its restriction to only positive tests is the Hitting Set Problem, dual to the Set Covering Problem (Aldridge 2019, Section 2.6), both of which are NP-complete (Karp 1972). For this reason we also consider a linear programming relaxation, widely used in previous work on group testing (Malioutov and Malyutov 2012; Aldridge 2019). The linear programming relaxation attempts to estimate ***x*** by replacing the constraint *x*_*j*_ ∈ [0, 1] with *x*_*j*_ ∈ [0, 1] for 1 ≤ *j* ≤ *N*. Since it is generally not binary, the solution of this linear program is rounded; for instance, any value greater than 0 may be replaced by 1. However, our experiments show that linear programming relaxation can increase the number of False Positives (FP), even though it provides faster computation times in some cases. We discuss this relaxation in more detail in Appendix A.

We also consider a formulation of the group testing problem that accounts for noise, similar to those appearing in (Zabeti, Dexter, Safari, Sedaghat, Libbrecht, and Chindelevitch 2020) based on previous work of (Malioutov and Malyutov 2012; Malioutov and Varshney 2013); see also (Aldridge 2019, Section 3.2). This formulation uses a vector of slack variables, denoted ***ξ***, to represent the possibility of a test result disagreeing with the one predicted from the status vector ***x***, and penalizes its components via two adjustable penalty coefficients *λ*_0_ > 0 and *λ*_1_ > 0. This leads to the following integer linear program:

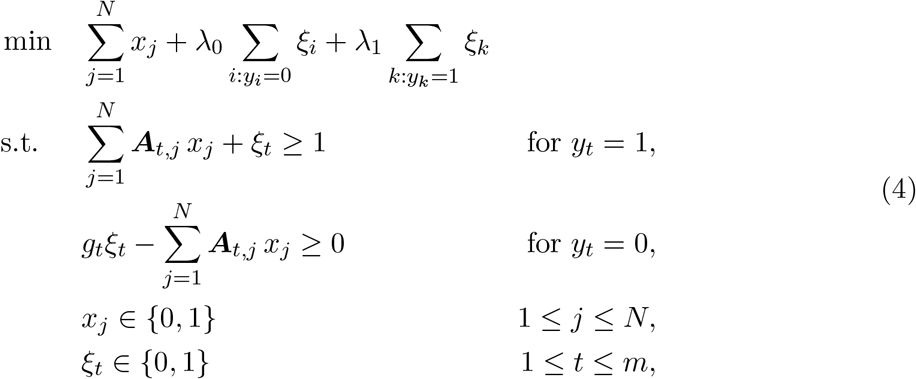

where 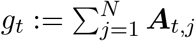 is the group size for group *t*.

In Section 3.1, we describe noise models relevant to the group testing problem. In the next section we discuss non-adaptive group testing schemes and the choice of the design matrix ***A***.

### 2.4. Non-adaptive group testing schemes

In non-adaptive group testing, the set of individuals being pooled in each test is determined in advance, independently of the outcome of every other test. This enables tests to be performed in parallel, a desirable feature for time-sensitive situations.

A key component of any non-adaptive group testing strategy is the design matrix. We focus on those designs that involve either a constant or near-constant divisibility *d* (defined as the number of groups each sample is present in), a constant or near-constant group size *g* (defined as the number of samples present in each group), or both. (Gebhard *et al*. 2020) studied unconstrained test designs and showed that random near-constant divisibility designs are information-theoretically optimal in the noiseless setting. They also studied bounded group size designs, and showed that the random near-constant group size design is optimal within this class of designs in the noiseless setting. A near-doubly-regular test design is one where both the divisibilities and the group sizes are near-constant. Random near-doubly-regular test designs, such as the one we consider in this work, have also been considered in (Mézard, Tarzia, and Toninelli 2008; Wadayama 2016) as part of a practical study of nonadaptive group testing. Our design differs from theirs in our selection of the group size based on an information-theoretical criterion – for details see Section 5.1.1. The algorithm described below has also been studied in (Miklós, Erdős, and Soukup 2013), where it was shown that the underlying Markov process performing edge swaps starting from an arbitrary realization allows for a uniform sampling of such near-doubly-regular test designs. Therefore the results presented in Section 5 are representative of the average performance of the algorithm over the range of prevalences and group sizes tested.

We also emphasize that the modular architecture of the **GroupTesting** package described in Section 4 allows for specification of alternative designs. For instance, the random near-constant divisibility design may be preferred when no group size constraints are imposed (Johnson, Aldridge, and Scarlett 2018), and explicit constructions based on disjunct matrices (Aldridge *et al*. 2019, §5.7), shifted transversals (Thierry-Mieg 2006) or low-density parity-check (LDPC) codes (Gallager 1962) can be substituted for the random near-doubly-regular design when deterministic designs are required.

#### Near-doubly-regular design construction details

We now describe the implementation of the random near-doubly-regular design. We use the **Python-igraph** package from https://igraph.org/python/ for this task, as described below.

We use the standard definitions, closely following (Wadayama 2016). Let *G* = (*V*_*L*_, *V*_*R*_, *E*) be a bipartite graph, called a *pooling graph*, such that *V*_*L*_ has *N* (left) nodes, *V*_*R*_ has *m* (right) nodes, and *E* is the set of undirected edges. We identify the nodes in *V*_*L*_ with the individuals, and the nodes in *V*_*R*_ with the group tests. We define an (*l, r, N*)-*regular pooling graph* as a bipartite graph *G* in which the left nodes have degree *l* and the right nodes have degree *r*. Since we wish to perform no more tests than in individual testing, we assume *m* ≤ *N* so that *l* ≤ *r*. A pooling graph represents the relationship between the individuals and the group tests they are part of, with an edge between *u* ∈ *V*_*L*_ and *v* ∈ *V*_*R*_ present if and only if the individual *u* is included in group test *v*.

Due to implementation details in **Python-igraph**, we rely on directed graph generation methods to generate random pooling graphs, with the convention that all edges have heads in *V*_*L*_ and tails in *V*_*R*_.

Given a pooling graph *G* with vertices *V*_*L*_ = {*v*_1_, …, *v*_*N*}_ and *V*_*R*_ = {*v*_*N*+1_, …, *v*_*N*+*m*_}and *V* := *V*_*L*_ *∪V*_*R*_, we define the *in-degree* of a vertex *v* ∈ *V*, denoted deg^*−*^(*v*), as the number of edges with head *v*, and similarly the *out-degree* of *v*, denoted deg^+^(*v*), is the number of edges with tail *v*. The in-and out-degrees of a directed graph must satisfy the *degree sum formula*:

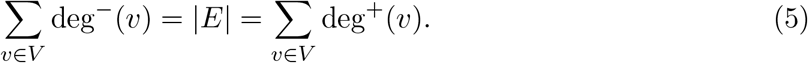

Here deg ^*−*^(*v*) for a sample *v* ∈ *V*_*L*_ is its divisibility, and deg^+^(*v*) for a group *v* ∈ *V*_*R*_ is its size. We define the *in-degree sequence* of *G* with vertices *V* = {*v*_1_, …, *v*_*N*+*m*_}as the sequence indeg(*G*) = (deg^*−*^(*v*_1_), …, deg^*−*^(*v*_*N*+*m*_)), and its *out-degree sequence* as the sequence outdeg(*G*) = (deg^+^(*v*_1_), …, deg^+^(*v*_*N*+*m*_)). If we require that the number of samples in each group *v* ∈ *V*_*R*_ is equal to *r* and the divisibility of each sample *v* ∈ *V*_*L*_ is equal to *l*, we can specify the desired pooling graph with the following in-degree and out-degree sequences:

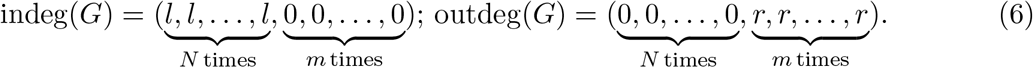

Note that these degree sequences ensure that the graph is bipartite. Figure 1 displays an example of pooling graphs generated with the **Python-igraph** package from in-degree and out-degree sequences specifying 8 groups (red) and 48 individuals (blue), where the groups have increasing size and corresponding individual divisibilities moving left to right.

**Figure 1:**
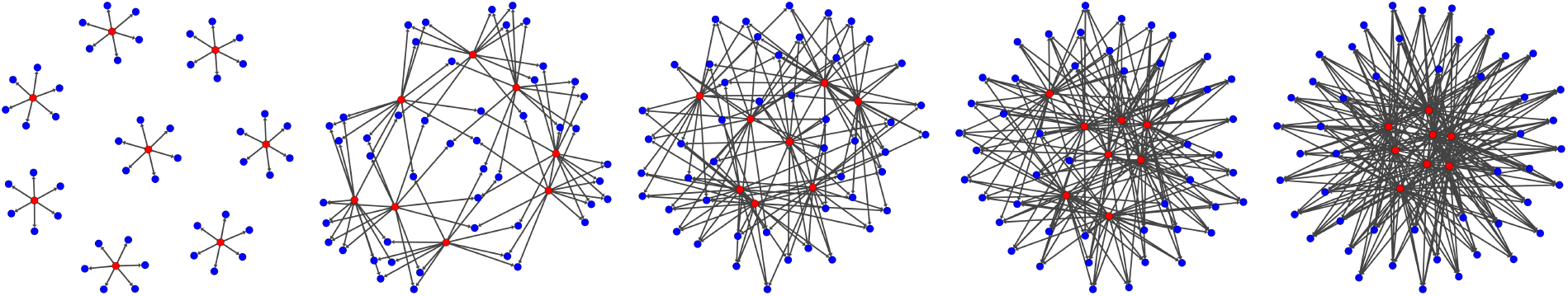
Example graphs generated with **Python-igraph** given *m* = 8, *N* = 48, and indeg(*G*) and outdeg(*G*) as in (6) with increasing *r* = 6, 12, 18, 24, 30 and corresponding *l* = 1, 2, 3, 4, 5 (left to right). Here the red nodes are groups and the blue nodes are individuals.

Note, however, that the degree sum formula (5) requires that *Nl* = *mr*, which may not be possible for some choices of parameters. In such a situation, we consider *constrained nearregular pooling graphs*, whose in-degree and out-degree sequences are of the form

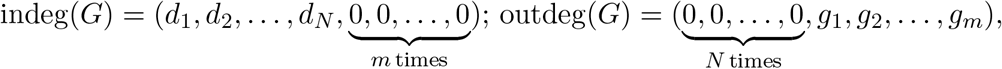

where each *g*_*t*_ = *g* to ensure that all the groups are full, each *d*_*j*_ ≤ *d*, and max_*k*_ *d*_*k*_*−* min_*l*_ *d*_*l*_ ≤1 to ensure that each sample has the same or almost the same divisibility. Since the quantity *mg/N* represents the average per-sample divisibility, we initially set

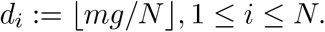

We then increase the divisibility of a random subset of *q* := *mg − Nd*_*i*_ of the samples, chosen without replacement, to *d*_*i*_ + 1 in order to satisfy (5); this is always possible since 0 ≤ *q < N*.

The Python-igraph interface allows graph generation from such degree sequences with multiple options. As the “`simple`” option can result in graphs with multiple parallel edges between a test *v* ∈ *V*_*R*_ and an individual *u* ∈ *V*_*L*_, we use the slower “`no_multiple`” option, which generates graphs without parallel edges. Figure 2 compares the outputs of these two options.

**Figure 2:**
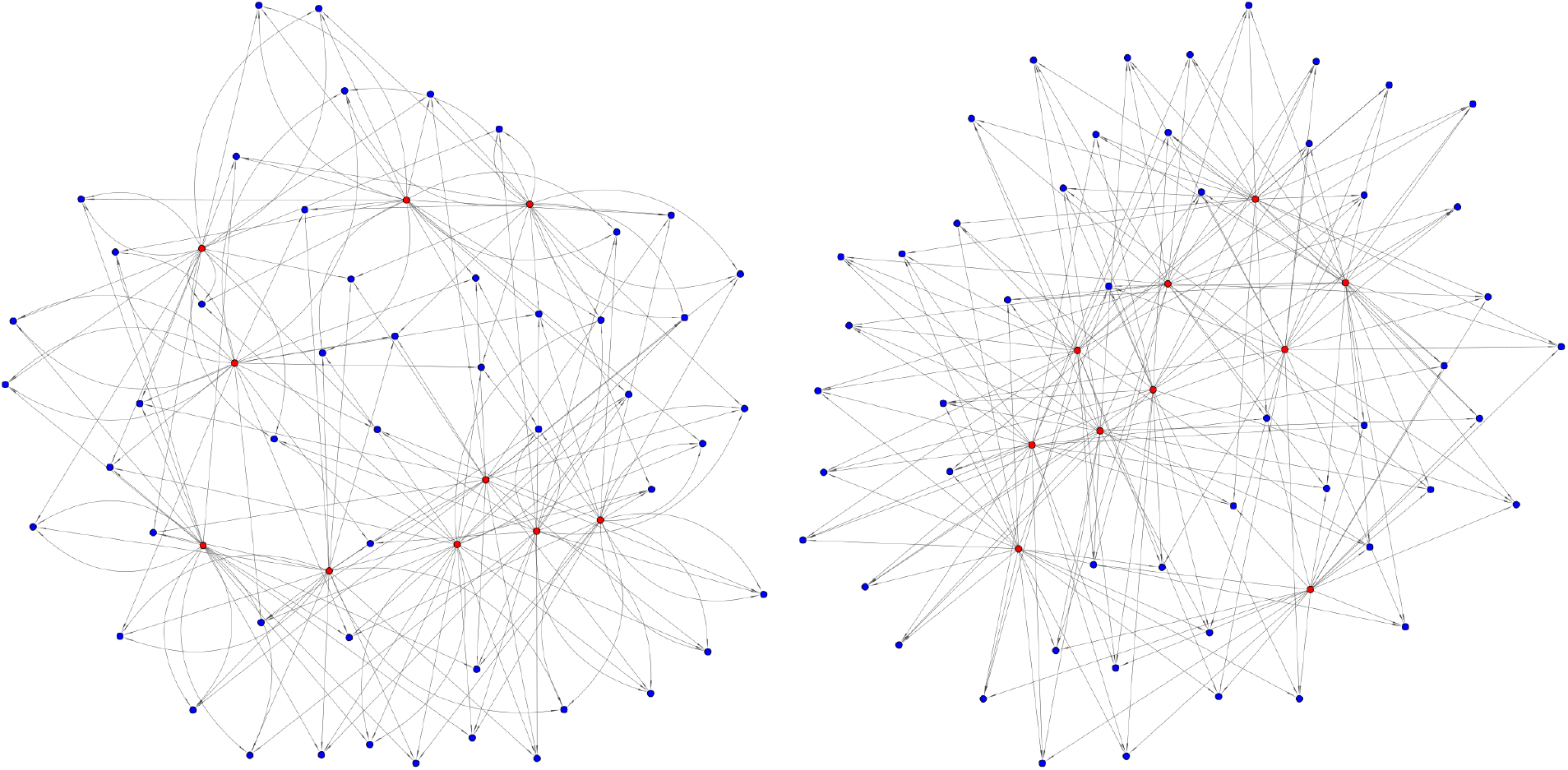
Comparison of “`simple`” (left) vs. “`no_multiples`” (right) graph generation methods in **Python-igraph**. See https://igraph.org/python/doc/igraph.GraphBase-class. html#Degree_Sequence for more details. Red nodes are groups, blue nodes are individuals.

## 3. Modelling SARS-CoV-2 testing in the laboratory

The primary method for testing for SARS-CoV-2 in the laboratory is via a polymerase chain reaction (PCR) test. This test seeks to amplify pieces of ribonucleic acid (RNA) that match a specific base-pair sequence, called a *primer*, designed complementary to the known genome sequence of the virus (Primerdesign 2020). As PCR tests directly detect the presence of the SARS-CoV-2 virus in an individual, they are considered to be the gold standard of testing, while other testing methods, such as antigen tests, may instead seek to ascertain previous exposure to the virus. Our main focus is on the PCR tests, and the constraints we impose are based on those, although most of the modules in our framework apply to other types of tests as well.

An individual PCR test can yield a false positive result (i.e. a positive result in a patient who is not infected) due to spurious RNA matches, which are rare, and the detection of viral RNA that is no longer part of actively replicating viruses, which is common (Pujadas *et al*. 2020). Similarly, an individual PCR test can yield a false negative result (i.e. a negative result in a patient who is infected) due to technical errors, which are rare, or the test occurring too soon after initial infection, which is common (Deeks, Dinnes, Takwoingi, Davenport, Spijker, Taylor-Phillips, Adriano, Beese, Dretzke, Ferrante di Ruffano *et al*. 2020).

The pooling of samples in the context of group testing can introduce additional errors of both kinds. Additional false positives may in principle occur when multiple negative samples are pooled together that each have a small amount of inactive viral RNA, but together combine to yield enough viral RNA to be detected by the test. Such false positives are highly unlikely due to the fact that each sample is diluted before being pooled (Primerdesign 2020), and will be disregarded in our error models. Additional false negatives, on the other hand, may result from the dilution effect, where the concentration of viral RNA in a positive sample gets reduced when it is combined with multiple negative samples. This effect is discussed in detail in several evaluation papers (Yelin *et al*. 2020; Shani-Narkiss *et al*. 2020; Hirotsu, Maejima, Shibusawa, Nagakubo, Hosaka, Amemiya, Sueki, Hayakawa, Mochizuki, and Omata 2020). We also revisit their results in the following section as we discuss the noise models implemented within our platform. Lastly, group testing is more prone to human error. Test tubes might be mislabeled in the test preparation, which is more likely to occur in group testing due to increased design complexity compared to individual testing. This can introduce both false positives and false negatives. We address the possibility of sample swapping as part of our discussion on noise models.

### 3.1. Noise models

To quantify the efficiency of group testing strategies and decoding in real-world scenarios, the sources of error discussed in the previous section must be accounted for through noise modeling. We now discuss some noise models relevant to the group testing setting that have been implemented in the **GroupTesting** package.

We assume that each test has the same probability distribution detebmining the outcome, and that the result of each group test is independent of all other group tests. We require the following definitions from (Aldridge *et al*. 2019) in defining noise models relevant to the group testing setting. We denote by ***𝒴*** the space of possible outcomes; for our purposes, ***𝒴*** = {0, 1}, where 0 indicates a negative test and 1, a positive test. For each outcome *y* ∈ ***𝒴***, *p*(*y*| *g, 𝓁*) is a probability distribution representing the probability of observing *y* from a group test of size *g* with *𝓁* infected individuals, and thus satisfies Σ_*y* ∈ ***𝒴***_ *p*(*y* | *g, 𝓁*) = 1. In the noiseless case

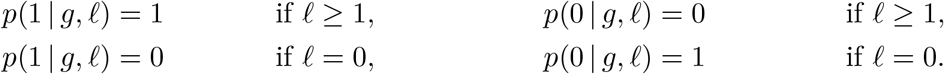

A simple noise model in which both false negatives and false positives can occur is the *binary symmetric* noise model. This has a parameter *ρ* ∈ (0, 1*/*2) and the probability distributions

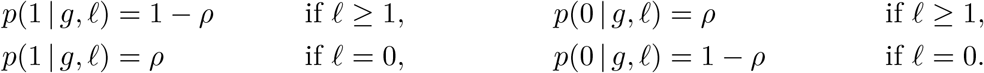

However, this model is often not realistic for the setting of group testing in the lab as false negatives and false positives typically occur with different probabilities. Two important noise models that are relevant to group testing in laboratory settings are *threshold* noise and *permutation* noise. Both models, in addition to the binary symmetric noise model included for completeness, are supported by the **GroupTesting** package. For usage details, see Section 4.

The threshold noise model is relevant to group testing problems where positive results are attained when the proportion of defective items in the test exceeds some upper threshold 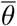, negative results are attained when the proportion is below a lower threshold *θ*, and false negatives occur at a rate of 1*/*2 when the proportion is between the two. A recent study by (Yelin *et al*. 2020) suggested that false negatives occur at a smaller rate than 1*/*2 in COVID-19 laboratory PCRtesting for the range of group sizes considered here. To account for the possibility of the false negative rate depending on the group size, in this work we consider a *generalized threshold model* in which false negatives occur at a rate *P*_FN_ ∈ (0, 1), specified as a parameter, when the proportion lies between *θ* and 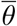. In this case, the probability distributions are given by

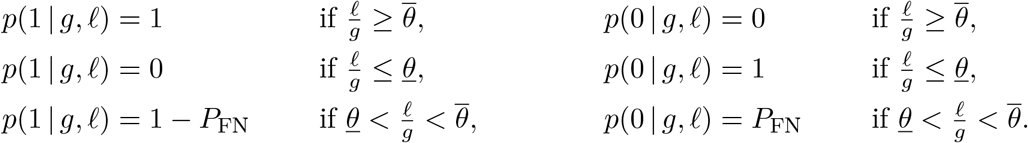

This model is relevant to testing scenarios where the laboratory testing process requires a minimum viral concentration *C*_min_ in a test for a positive test result. In groups of size *g* each positive sample contributes *C* sample≥ *C* min */g*. A false negative result occurs if the total concentration of all *𝓁* positive samples 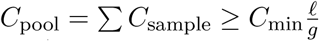 is smaller than *C*_min_, where the inequality explains the scaling with *𝓁/g*.

Permutation noise occurs when the test results themselves are accidentally shuffled or mislabeled, and is a serious source of error that must be accounted for. In this case a small number of pairwise swaps of the components of the test outcome vector ***y*** may occur, leading to some test results no longer being correlated with their assigned group. In realistic laboratory testing scenarios, only a small number of test results will be swapped. These effects can be modeled by a discrete probability distribution such as the negative binomial or the geometric distributions. The **GroupTesting** package supports these types of studies by allowing for a user-supplied value *ρ* ∈ [0, 0.5] as input specifying the proportion of tests to be permuted. The code then picks uniformly at random two items to swap until *k* = ⌈*ρm*⌉ pairwise swaps (without replacement) have been performed. Since these types of errors are common in high-throughput testing scenarios requiring human intervention, group testing and decoding procedures must be robust to these human errors.

## 4. Software for solving group testing problem

In this section we describe our main contribution, a `Python` (van Rossum 1995) software package named **GroupTesting** which enables rigorous evaluation of the efficiency of group testing schemes and solvers over a range of parameterizations. The **GroupTesting** package is implemented in `Python`, and is freely available on GitHub at https://github.com/WGS-TB/ GroupTesting and through PyPi under **GroupTesting**. The package follows a modular design, allowing individual substitution and analysis of each component, and is focused on reproducibility and ease of configuration. Figure 3 displays a diagram of the core modules of the package and a typical workflow.

**Figure 3:**
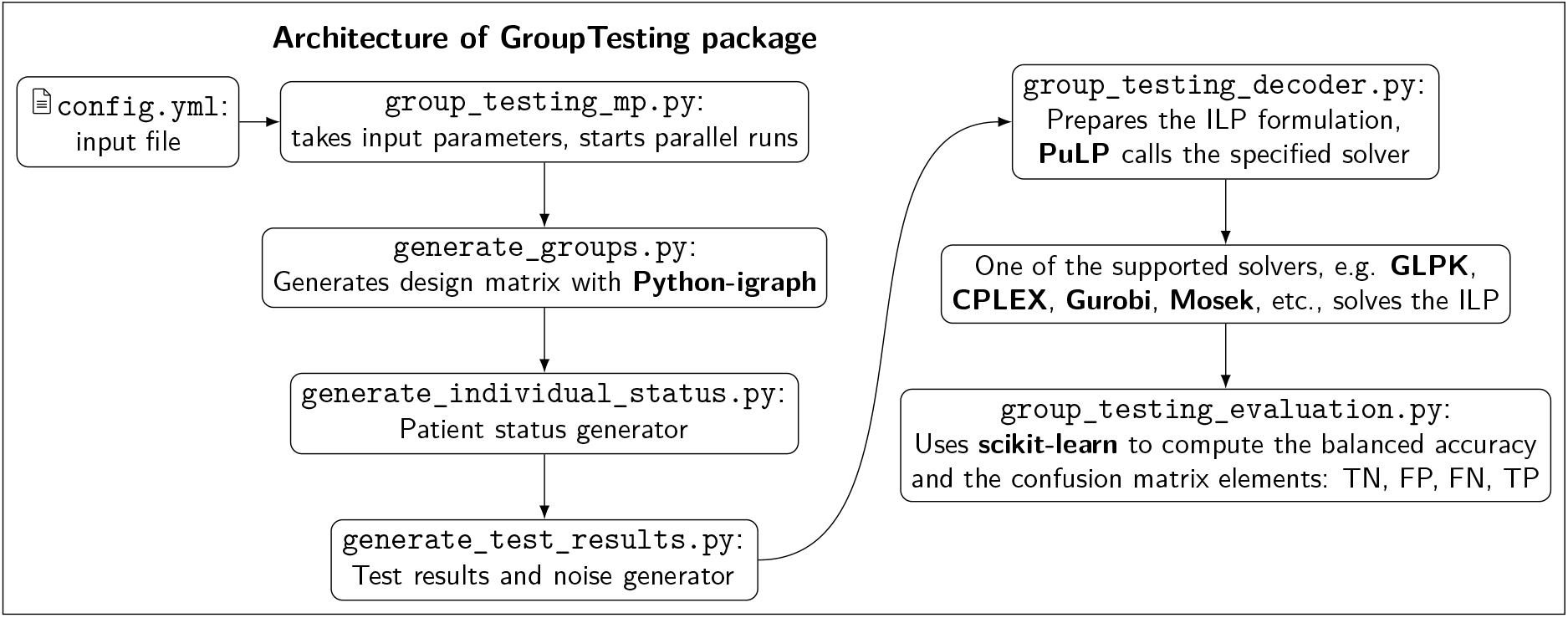
Modular design of the **GroupTesting** framework.

`config.yml` is the control center of the framework. It allows the user to build a group testing pipeline, specify each core module that is needed, and parameterize that module. Listing 1 describes the format of the configuration file – a complete tutorial is available on the GitHub repository.

`config.yml` is divided into two main dictionaries, `design` and `decode`, representing the parameters of the design and decoding stages of the framework, respectively. `design` contains three sub-dictionaries: `groups`, `individual_status` and `test_results`, corresponding to the core modules `generate_groups`, `generate_individual_status`, `generate_test_results` respectively. `groups` (Listing 1, lines 2-8) specifies the features of the design matrix **A** such as its dimension (the population size *N* and the number of tests *m*), the group size *g*, and the divisibility *d*. `generate_groups.py` uses these parameters to generate the desired design matrix.

**Listing 1:**
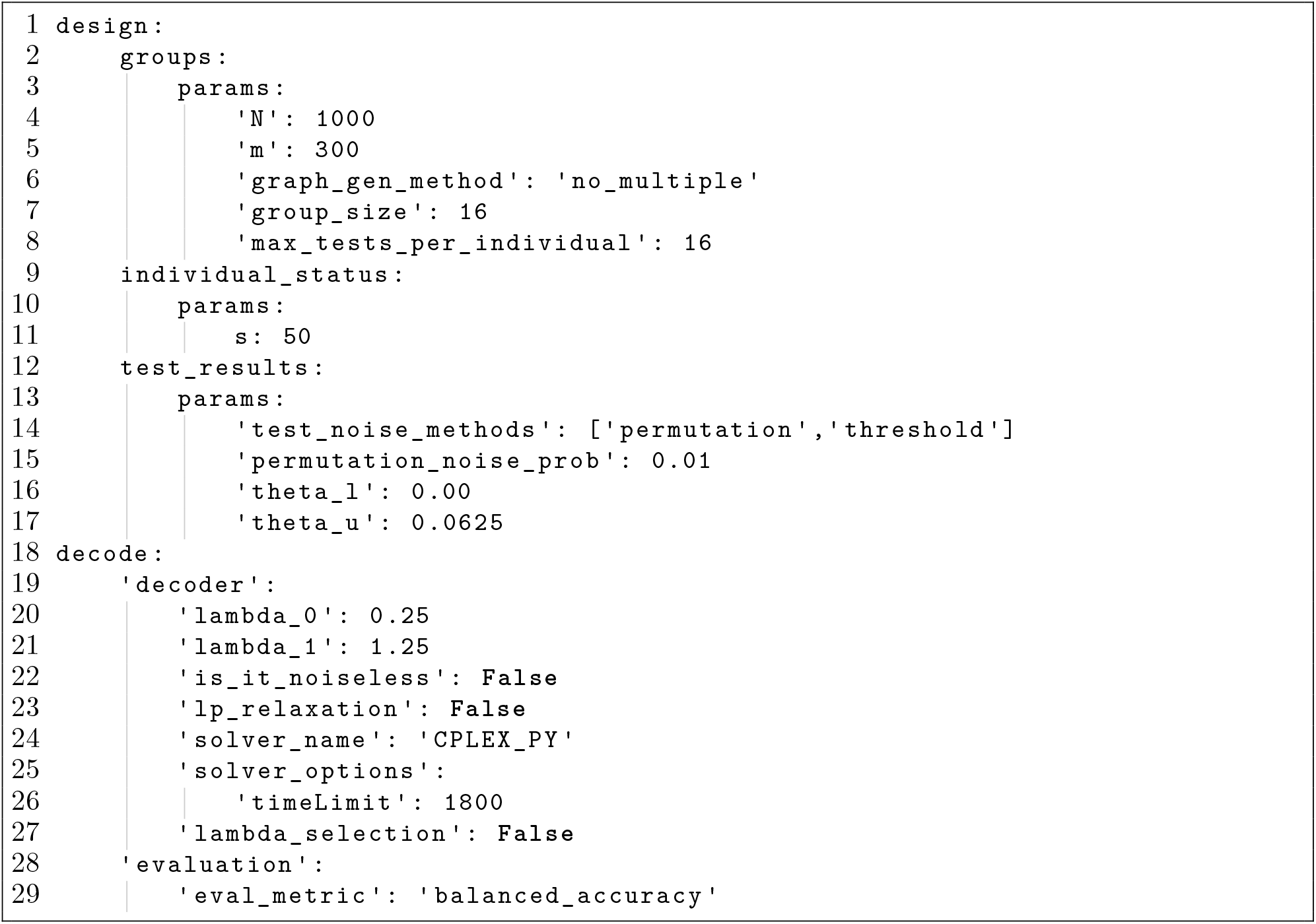
General format of `config.yml file`.

`individual_status` (Listing 1, lines 9-11) records the number of infected individuals in the population. This parameter is then used by `generate_individual_status.py` to randomly assign an infected status to *K* individuals in the population. The remaining individuals are labeled as not infected. This assignment is represented by the binary status vector.

`test_results` (Listing 1, lines 12-17) contains information relevant to the noise models for the given testing scenario. This can be specified by selecting one or more models from the list of available model options (‘threshold’, ‘binary_symmetric’, ‘permutation’). These models are then applied to generate the results of the group tests via the `generate_test_results.py` module.

Similar to `design, decode` (Listing 1, lines 18-29) contains two sub-dictionaries, `decoder` and evaluation, which represent the parameters corresponding to the two core modules, respectively named `group_testing_decoder` and `group_testing_evaluation`.

`decoder` (Listing 1, lines 19-27) contains all the decoder’s arguments. Since we employ the **PuLP** (Mitchell, O’Sullivan, and Dunning 2011; Lougee-Heimer 2003) package to set up the ILPs (4), (3) and their LP relaxations, this sub-dictionary can also contain **PuLP** parameters such as the solver name and solver options. Users can pass different parameters to their chosen solver, as long as they are allowable by the chosen solver and **PuLP**, through the `solver_options` sub-dictionary. The core module `group_testing_decoder.py` is then responsible for setting up the ILP problem. This module uses the **PuLP** package to specify the ILP’s variables, objective function, and constraints as expressions, and interfaces with the supported solvers to compute a solution. See Section 4.2 for more details on the supported solvers and examples of problem formulation.

This module is also able to perform cross-validation to select the parameters *λ*_0_ and *λ*_1_ when the objective is to solve the problem (4). The cross-validation is performed using `GridSearchCV` in **scikit-learn** (Pedregosa, Varoquaux, Gramfort, Michel, Thirion, Grisel, Blondel, Prettenhofer, Weiss, Dubourg *et al*. 2011). To do so, Listing 1 should be modified as follows:

**Listing 2:**
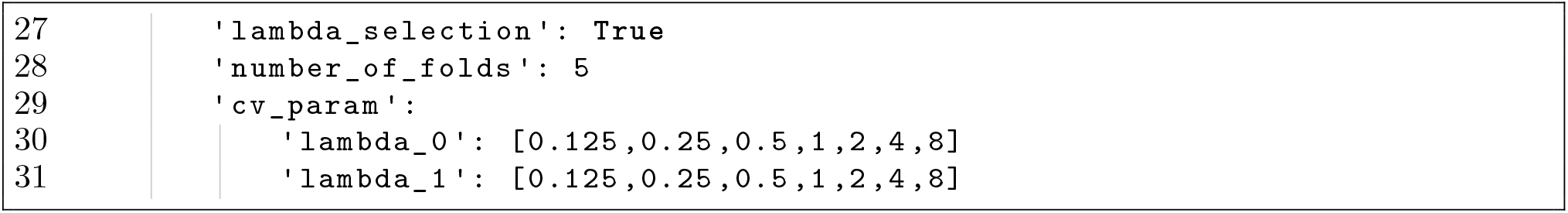
Example of selecting the parameter *λ*_0_ and *λ*_1_.

The framework also allows the user to set *λ*_0_ = *λ*_1_ when performing cross-validation. This setting can save a considerable amount of computing time, and is relevant for experiments in a noise model with a symmetric error probability, or when the user would like the ILP (4) to penalize the false positives and the false negatives equally. Listing 3 demonstrate this option, with *λ*_*e*_ = *λ*_0_ = *λ*_1_, where subscript *e* stands for equal.

`evaluation` (Listing 1, lines 28-29) contains evaluation metrics. After decoding is complete, the results can be compared to the true solution using the `group_testing_evaluation.py` module. This module provides the confusion matrix along with the desired evaluation metric such as the balanced accuracy, the sensitivity, or the specificity using **scikit-learn**.

**Listing 3:**
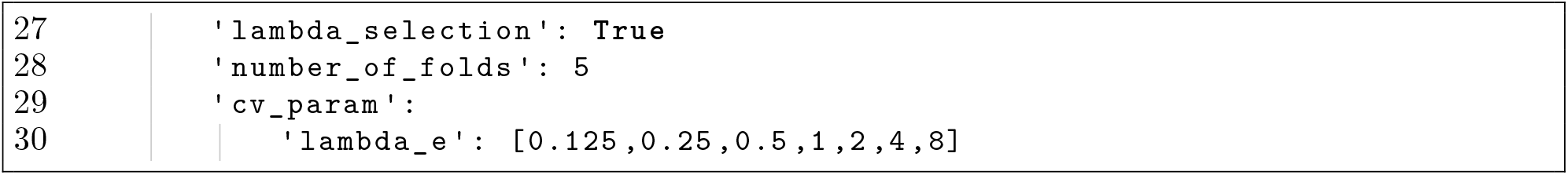
Example of selecting the parameter *λ*_*e*_.

**Listing 4:**
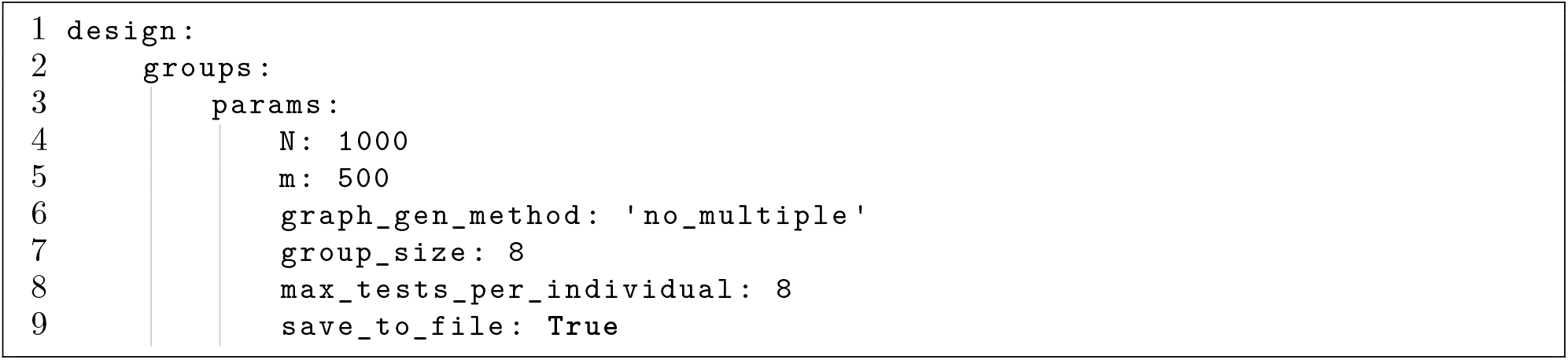
Example of when only generating a design matrix is needed.

After modifying the configuration file, the package can be run from the command line via:
*GroupTesting --config=PATH_TO_CONFIG*.*yml*

### 4.1. Customization and performing series of experiments

The **GroupTesting** framework is modular and customizable to the users’ needs. In the following section we provide different possible use cases. In the first one, only the design matrix is being varied. In this case the user only needs to define the `groups` block. Listing 4 is a example of a complete `config.yml` file for this scenario.

Another possible use case is a pre-defined design matrix loaded from a comma-separated values (CSV) file. Instead of providing a list of parameters, the user only needs to provide the filename, as shown in Listing 5. This feature is also available for the next two modules, `individual_status` and `test_results`. Note that when an input file is provided for a block, no parameters need to be specified, as their values are automatically extracted from the input file.

Users are also able to provide their own module to substitute any with of the core-modules in Figure 3. This feature is accessible by the `alternative_module` keyword, followed by the name of the module, the name of the function needed to operate it, and a list of its parameters. Listing 6 demonstrates how to get the framework to use a custom module to generate test results. In this example, `custom_test_results` is the name of the module, `test_vector_generator` is the function name, and `p1`, `p2` and `p3` are the argument values to be passed to the function.

**Listing 5:**
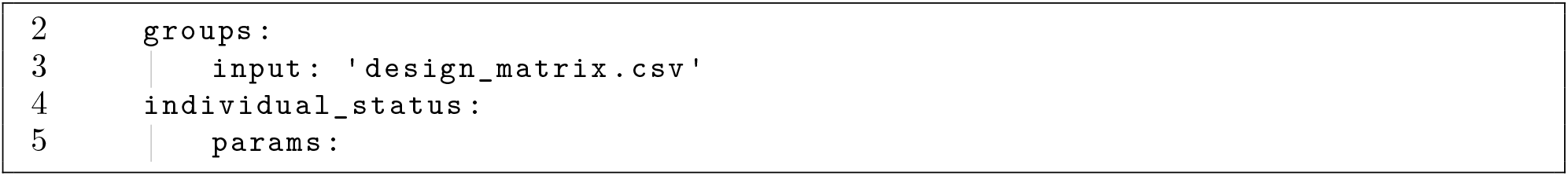
The `groups` dictionary accepts a pre-defined design matrix as an input.

**Listing 6:**
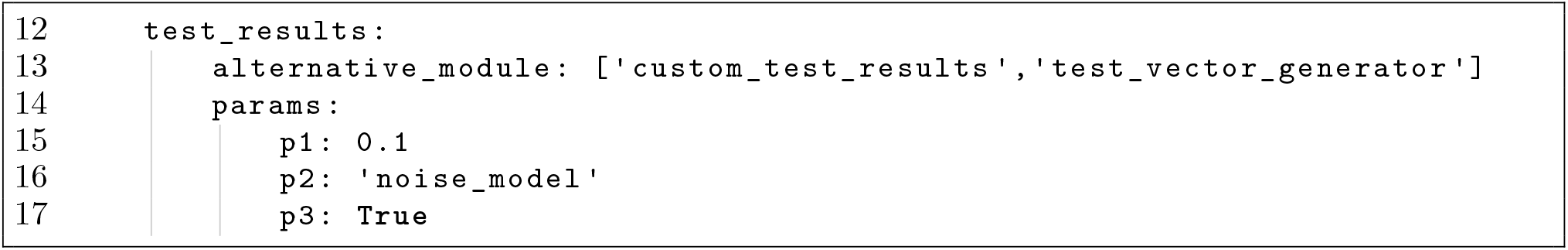
Generating the test results with the `custom_test_results.py` module.

Moreover, the framework allows the user to perform a series of experiments with a single `config.yml file`. For example, if an experiment similar to Listing 1 is to be performed, but the user would like to compare the performance of group testing for group sizes 4, 8, and 16 on a different range of infected individuals, from *K* = 50 up to, but not including, *K* = 100, with a step size of 10, this can be achieved as follows:

**Listing 7:**
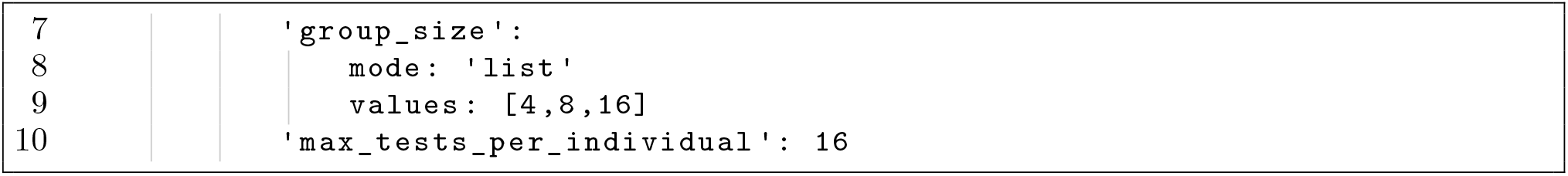
`config.yml` accepts a list of values for a parameter.

**Listing 8:**
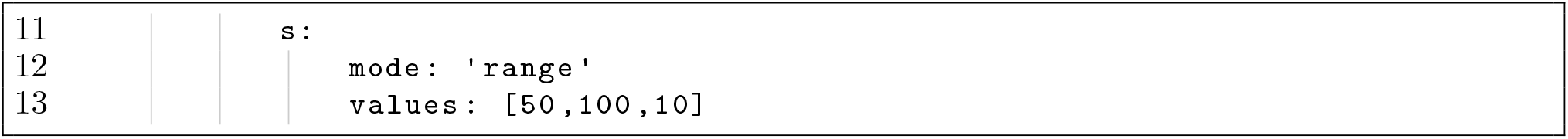
`config.yml` accepts a range of values for a parameter.

In this case, the framework performs a set of parallel experiments based on all possible combinations of the parameters in `config.yml`, by distributing the corresponding experiments over the available processors. The framework uses the Pool object of the **multiprocessing** package (McKerns, Strand, Sullivan, Fang, and Aivazis 2012) for this purpose. Note that the framework does not support nested parallelization, so if a series of experiments also contains cross-validation for selecting parameters *λ*_0_ and *λ*_1_, a process that is also performed in parallel, the framework automatically performs the experiments sequentially while executing each cross-validation in parallel.

### 4.2. Optimizers

The **GroupTesting** package supports a variety of solvers through the **PuLP** application programming interface (API). Instead of specifying the ILP problem through input files, e.g. mathematical programming system files, we rely on the **PuLP** API to directly specify the problem. The **PuLP** package supports a variety of popular open-source and commercial solvers, including pkgCBC, **CHOCO, COIN, CPLEX, MIPCL, GLPK, Gurobi, Mosek**, and **SCIP**.

Furthermore, the config file makes it convenient to customize the decoding ILPs. For instance, we can specify whether our problem is noiseless (i.e. ILP (3)) or noisy (i.e. ILP (4)) by setting the `is_it_noiseless` parameter to `True` or `False`, respectively. Similarly, to solve the ILP or replace it with an LP (as described in Appendix A), we can use the Boolean parameter `lp_relaxation`. The solver can be specified by `solver_name` and the solver’s arguments can be set by `solver_options`. Listing 1, lines 19-27, provides an example of noisy group testing without LP relaxation, while Listing 9 demonstrates a noiseless group testing problem with LP relaxation.

**Listing 9:**
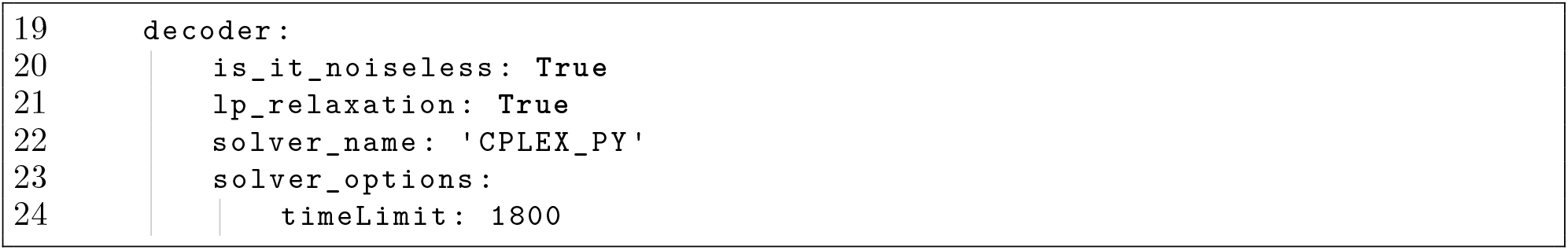
Example of noiseless group testing with LP relaxation.

## 5. Numerical experiments

In this section we include a selection of numerical experiments exploring the efficiency of the **GroupTesting** package at identifying infected individuals from large populations. We study the problem posed in Section 2.3 with the non-adaptive group testing scheme based on the doubly-regular design from 2.4 in both the noiseless setting and in the presence of small amounts of thresholding and permutation noise, as described in Section 3.1. We parameterize and solve the resulting problems using the **PuLP** package with the **CPLEX** solver, see Section 4, testing the efficacy of the proposed approach over a range of group sizes. Finally, we record our results in terms of the balanced accuracy, averaged over 10 trials.

### 5.1. Evaluation metrics

#### Effect of group size

A major question in practical group testing is how to set the maximum group size *g* given an estimate for the prevalence *p*. From an information-theoretic perspective, it is desirable that each test yields a positive result with a probability 1*/*2. In the noiseless setting, the probability that a group yields a negative result is (1 *− p*)^*g*^. Hence, equating this with 1/2 leads to the following selection rule for *g*:

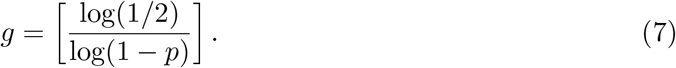

Here [*x*] denotes the nearest integer to *x*.

The first set of experiments we conduct aims to empirically study the effect of the group size on the average balanced accuracy, given the same non-adaptive testing strategy and budget of group tests *m*. We conduct this experiment in the noiseless setting with a population of size *N* = 1000, studying the efficacy of the proposed approach in recovering the status vector ***x*** by solving problem (4) with group size *g* ∈ {4, 8, 16, 32} and increasing *m* up to *N*.

Figure 4 displays the results of solving this problem at various prevalences *p*. In Table 3, we compare the information-theoretic group size (7) to the empirical results. We note that the best-performing group sizes from the experiments match the information-theoretic choice for larger prevalences. For smaller prevalences, namely, 0.5% and 1.0%, the information-theoretic group size is much larger and beyond the maximum allowed group size of 32 considered reasonable in this study (see Section 2.1); however, in this regime group sizes at or just below this allowed maximum exhibit very good performance, providing near perfect recovery after approximately 100-150 tests in the case *g* = 32 and 200-250 tests when *g* = 16. In contrast over 450 tests are required for a prevalence of 0.5% when *g* = 4.

**Table 3:** Recommended group sizes based on information theory (7) and empirical observations of solutions to (3) with a population of size *N* = 1000, see Fig. 4. The last two rows show the percentage of tests saved relative to individual testing that allows a balanced accuracy above 0.90 and 0.95, respectively.

| Prevalence $p$ | 0.5% | 1.0% | 5.0% | 10.0% | 15.0% | 20.0% |
| --- | --- | --- | --- | --- | --- | --- |
| Information-theoretical group size (7) | 138 | 69 | 14 | 7 | 4 | 3 |
| Empirical best group size | 32 | 32 | 16 | 8 | 4 | 4 |
| Percentage saving (balanced accuracy 0.90) | 92% | 90% | 70% | 50% | 34% | 22% |
| Percentage saving (balanced accuracy 0.95) | 91% | 89% | 65% | 40% | 20% | 5% |

**Figure 4:**
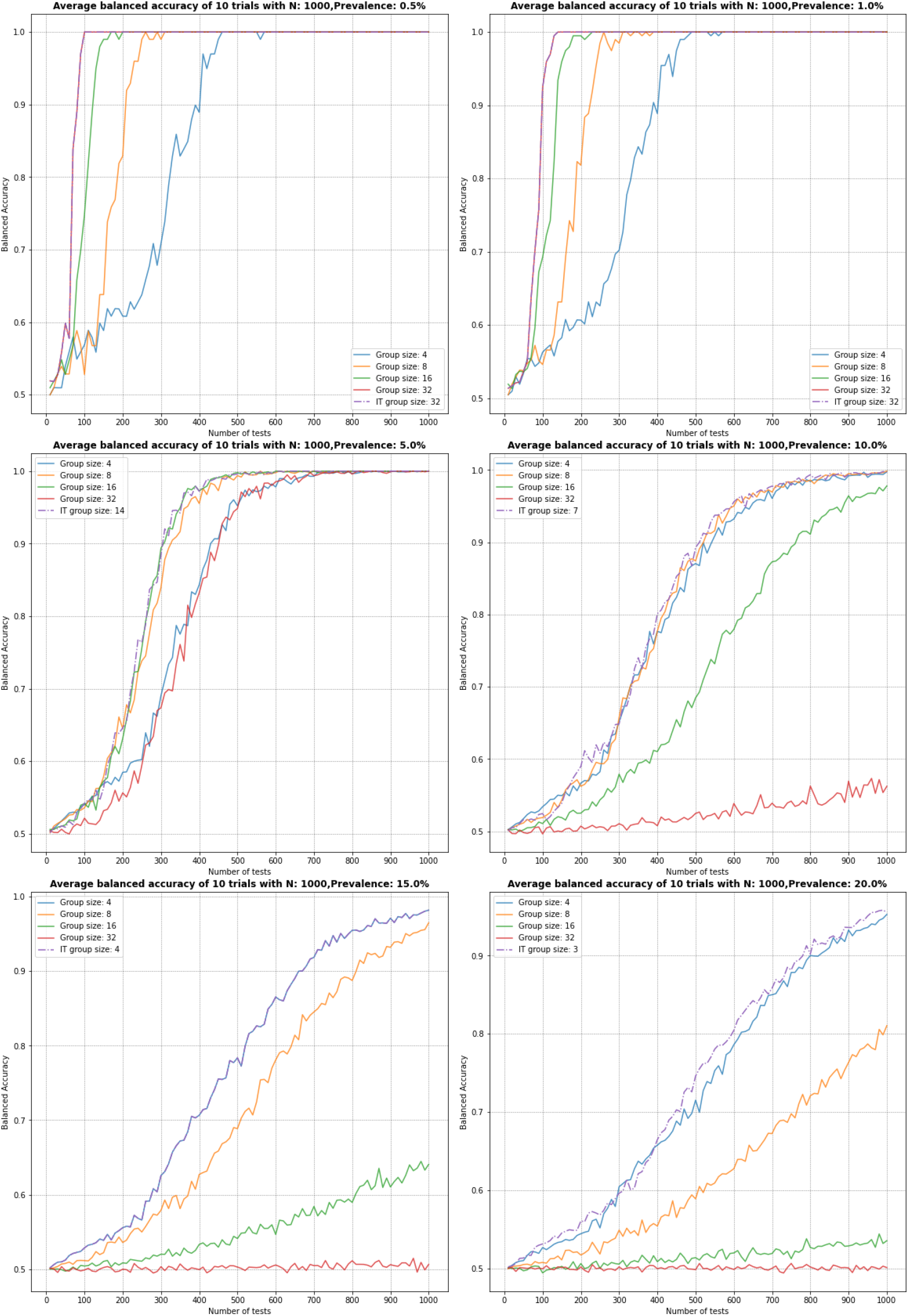
Comparison of the average balanced accuracy over 10 trials across a range of group sizes from 4 to 32 and Information Theoretical (IT) group size, for prevalences from 0.5% to 20%.

However, as the prevalence increases, the efficiency of larger group sizes dramatically decreases. For *p* = 5% and group size *g* = 32, over 700 tests are required for near-perfect recovery, and for the larger prevalences tested (namely 10%, 15% and 20%) this strategy never achieves a balanced accuracy above 0.6. This behaviour is expected, since when the prevalence is higher, larger group sizes yield tests that predominantly return positive values. For the smaller group sizes, we also observe a loss of accuracy as the prevalence increases, and at a 20% prevalence we require over 800 samples for a balanced accuracy of 0.9. In the high prevalence regime, the proposed strategy of non-adaptive group testing with the doubly-regular design conveys far less benefit over individual testing of the entire population. We leave a study of the efficacy of alternative designs in this setting to future work, although the performance of any group testing design is expecting to decrease at such a high prevalence.

In Table 3 we report the percentage saving offered by the best-performing group testing scheme over fully testing the entire population. For small prevalences (0.5% and 1.0%), group testing with the best-performing group size achieves over 90% savings. Moreover, the savings remain above 50% even for prevalences as large as 10%.

#### Large population test

Figure 5 displays the result of solving the group testing problem with a population size of *N* = 10, 000, for information theoretically optimal group sizes and various prevalences. For small prevalences we see near-perfect recovery after approximately 1500 group tests, suggesting a savings of 85% over fully testing the entire population. These results are similar to the ones with *N* = 1000, suggesting that the near-doubly-regular test design is well-suited for tackling such large population sizes.

**Figure 5:**
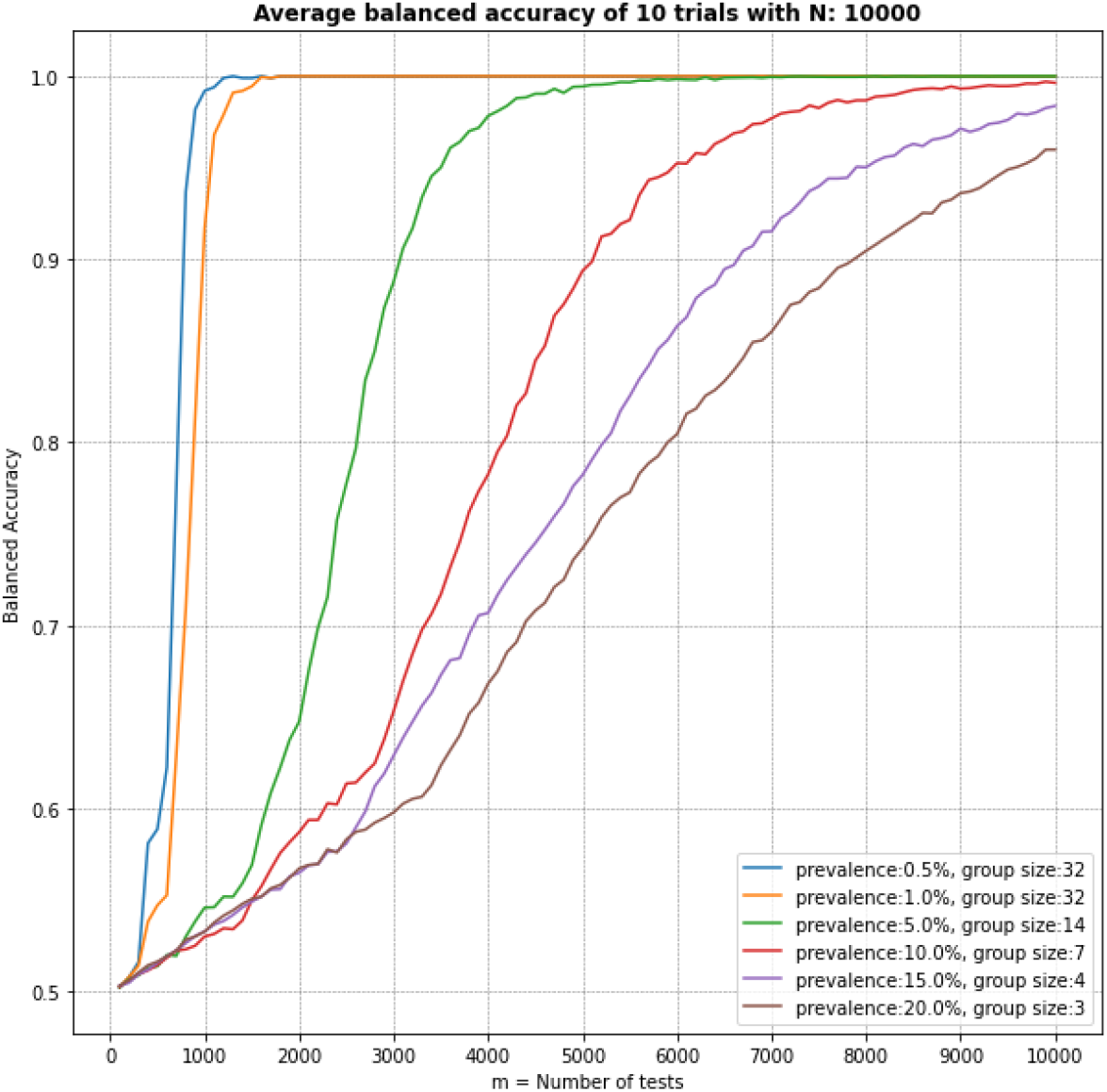
Comparison of average balanced accuracy over 10 trials with *N* = 10000 using the information theoretical group size, for prevalences from 0.5% to 20%.

#### Noisy experiments

To examine the effect of noise on the performance of the solvers under realistic laboratory testing scenarios, we include a selection of numerical results with both the threshold noise model, with parameters chosen based on observations from (Yelin *et al*. 2020), and the permutation noise model with various permutation levels *ρ*. We solve problem (4) along with using cross-validation for selecting *λ*_0_ and *λ*_1_ where *λ*_0_ = *λ*_1_ = *λ*_*e*_. The **GroupTesting** platform provides the facility for performing cross-validation, either on the single parameter *λ*_*e*_ or on both parameters *λ*_0_ and *λ*_1_, which we describe in more detail in appendix B.

In (Yelin *et al*. 2020) it was observed that false negatives occur at a rate of approximately 10% when a group of size *g* ≥16 contained one infected individual, and did not occur for lower dilutions. The effect of these levels of false negative rates can be modelled by threshold noise with parameters *θ* = 0 and 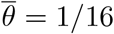 and probabilities

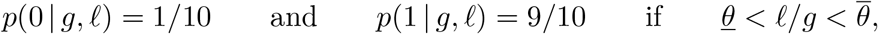

corresponding to *P*_FN_ = 1*/*10 in the model from Section 3.1. We study the effect of this noise model with group testing with groups of size *g* = 32, a population of *N* = 1000 individuals, and a range of prevalences *p* ∈ {0.5%, 1%, 5%}in Figure 6. There we can see that for small prevalences, e.g., 0.5% or 1%, to achieve near-perfect recovery the solvers require approximately 50% more samples over the noiseless case. However at a prevalence of 5% the solvers require approximately 700 samples to achieve near-perfect recovery in the noiseless case, but with threshold noise the maximum of the averaged balanced accuracy scores over all values *m* is only 0.987 even with *m* = 1000 group tests.

**Figure 6:**
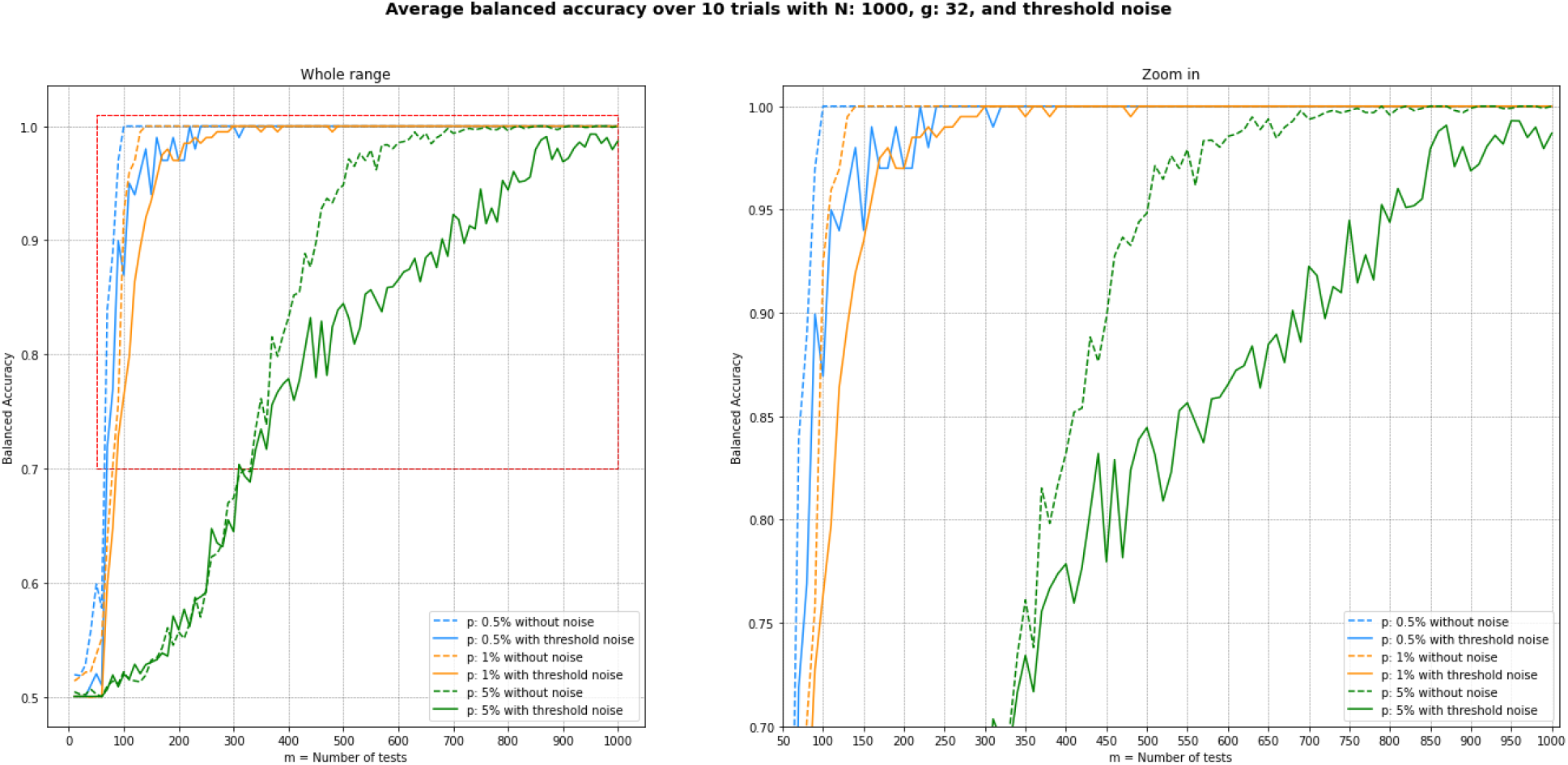
Comparison of average balanced accuracy with and without presence of threshold noise over 10 trials with *N* = 1000, *g* = 32 across a range of prevalence from 0.5% to 5%

Our permutation noise experiments aim to examine the effect of small amounts of random mislabeling that may occur during testing. Figure 7 shows the results obtained with group size *g* = 16, prevalence *p* = 5%, and values of *ρ* from 1% to 5%. The results indicate that small amounts of permutation noise marginally lower the effectiveness of the solvers, decreasing proportionally with the noise level. Nonetheless, with all noise levels considered near-perfect recovery was obtained after approximately 600-650 samples, as in the noiseless case.

**Figure 7:**
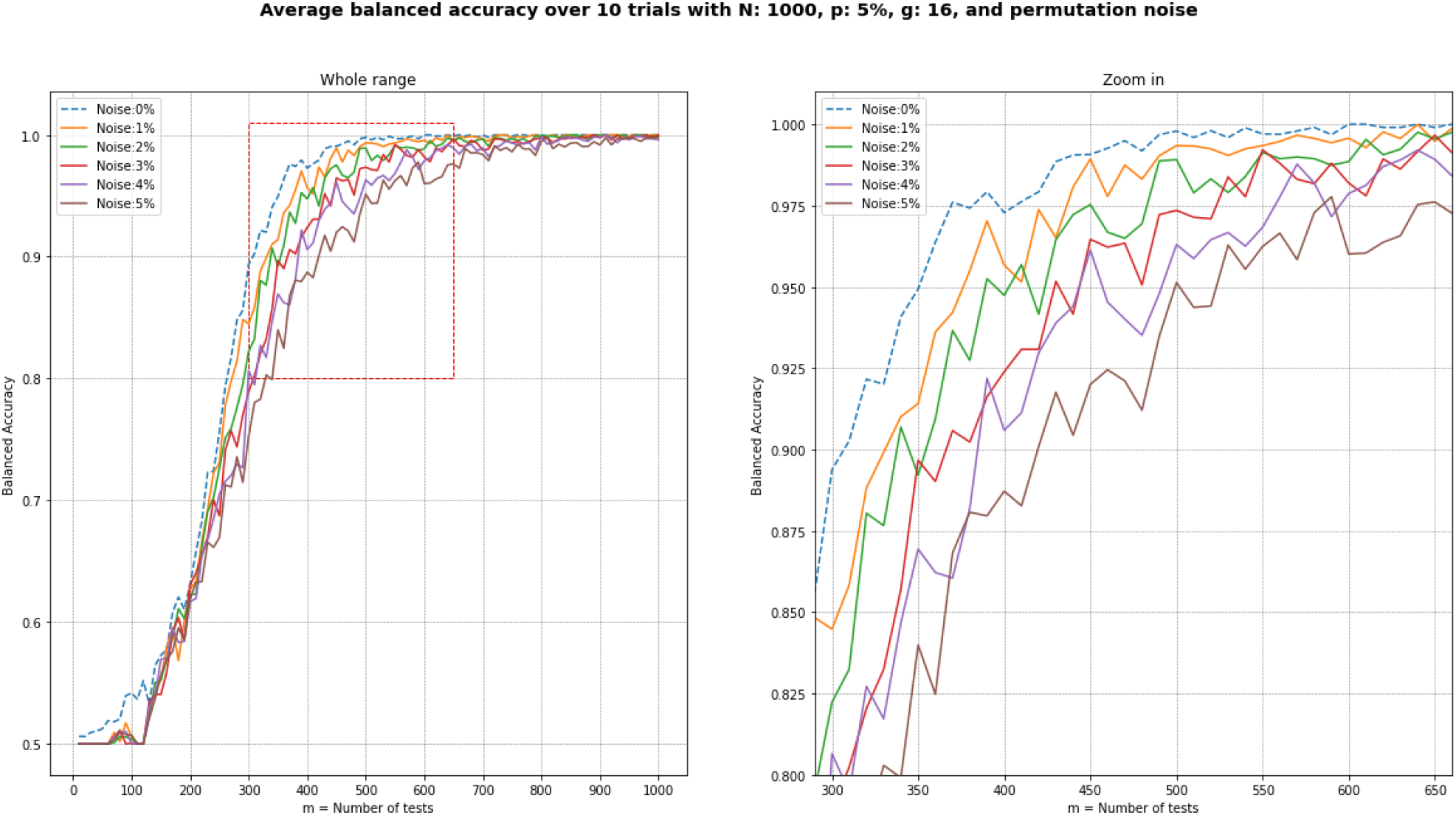
**Left:** Comparison of average balanced accuracy over 10 trials with *N* = 1000, *g* = 16 and *p* = 5% across a range of permutation noise from 0% to 5%, **right:** same figure zoomed in.

## 6. Summary and discussion

This work has focused on estimating whether or not an individual is infected based on group tests that return only positive or negative values for each group. As discussed in Section 2.2, another line of recent work aims to compute the individual viral load, given viral load data from each test (Ghosh *et al*. 2020b). This may not be realistic in standard COVID-19 testing. However, a variation is to consider the *cycle threshold* data for each group. The cycle threshold is closely related to the viral load, but unlike the latter, can be measured in practical COVID-19 tests. Using this data to infer cycle threshold values for each individual may not be desirable in practical settings, due to concerns about how these values may be interpreted by individuals (Rhoads, Peaper, She, Nolte, Wojewoda, Anderson, and Pritt 2020). However, one could use this data solely to estimate whether an individual is infected or not. We anticipate that this approach may lead to non-negligible improvements in the efficacy of group testing, and intend to pursue this in future work.

An additional direction previously explored in the literature has focused on using group testing to estimate only the prevalence of infection in a population, rather than identifying the infected individuals. Under plausible assumptions, this can not only require fewer tests (Tu, Litvak, and Pagano 1994), but also provide more confident estimates (Petito and Jewell 2016). The two approaches can also be combined; first, a certain number of group tests can be performed to estimate the prevalence, and then, a group testing design can be selected based on the estimated prevalence. However, our framework focuses on the case where the prevalence is known, at least approximately.

In conclusion, our framework provides an open-source robust and reproducible platform for evaluating the effectiveness of non-adaptive group testing approaches, with a particular emphasis on SARS-CoV-2 testing via PCR tests. Thanks to its modular structure, it allows for the exploration of a large number of parameters, designs, decoding algorithms, noise models and evaluation metrics. By leveraging **PuLP** it also allows the comparison of different solvers for linear and integer linear programs. It is our hope that it will provide an impetus for further exploration of this paradigm, both in theory and in clinical and public health practice.

## Data Availability

All data is generate via the pipeline introduced in the manuscript.

https://github.com/WGS-TB/GroupTesting

## Acknowledgments

This project was partially funded by a Genome Canada grant, “Machine learning to predict drug resistance in pathogenic bacteria”. LC acknowledges funding from the MRC Centre for Global Infectious Disease Analysis (MR/R015600/1), funded by the UK Medical Research Council (MRC) and the UK Foreign, Commonwealth & Development Office (FCDO) under the MRC/FCDO Concordat agreement, and is part of the EDCTP2 program supported by the EU. BA acknowledges funding from NSERC through grant R611675. BA and ND acknowledge support from PIMS (Pacific Institute for the Mathematical Sciences), through the CRG “High Dimensional Data Analysis”.

## A. Performance of ILP versus LP

In this section, we include a comparison of the results when solving the ILP (3) versus solving its LP relaxation, defined as follows:

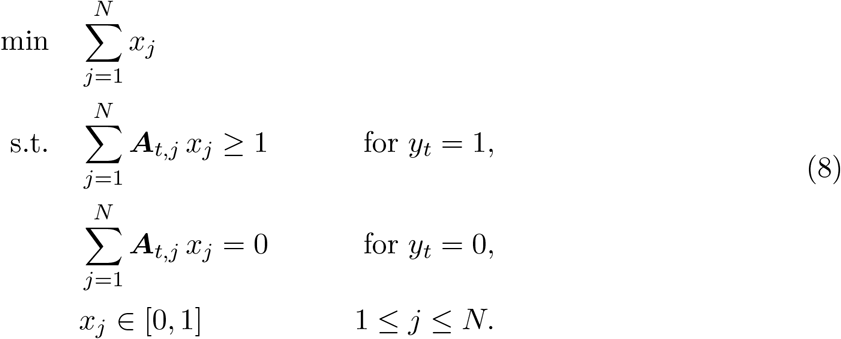

Note that in the latter, the constraint *x*_*j*_ ∈ {0, 1} is relaxed to 0 ≤ *x*_*j*_ ≤ 1 to give an LP. Once this program is solved, the solution is rounded by replacing any value greater than a given threshold parameter *β* by 1 and any value less than *β* by zero. In this study, we use the threshold value *β* = 0. Figure 8 gives a comparison of the averages over 10 trials of the balanced accuracy, sensitivity, specificity, and system time (measured in ticks) to solve when *N* = 1000, *g* = 32 and *p* = 0.5% or *p* = 1.0%. We also include the plot ranges covering the minimum and maximum of these statistics taken over all trials. There we can see a decrease in the specificity of the LP approach near the point when the balanced accuracy of both methods crosses 0.75 (approximately *m* = 60 tests when *p* = 0.5% and *m* = 80 tests when *p* = 1%). We also note a sharp increase in the system time to solve for the ILP near these points, suggesting that the solvers required more iterations on average to find a suitable solution.

**Figure 8:**
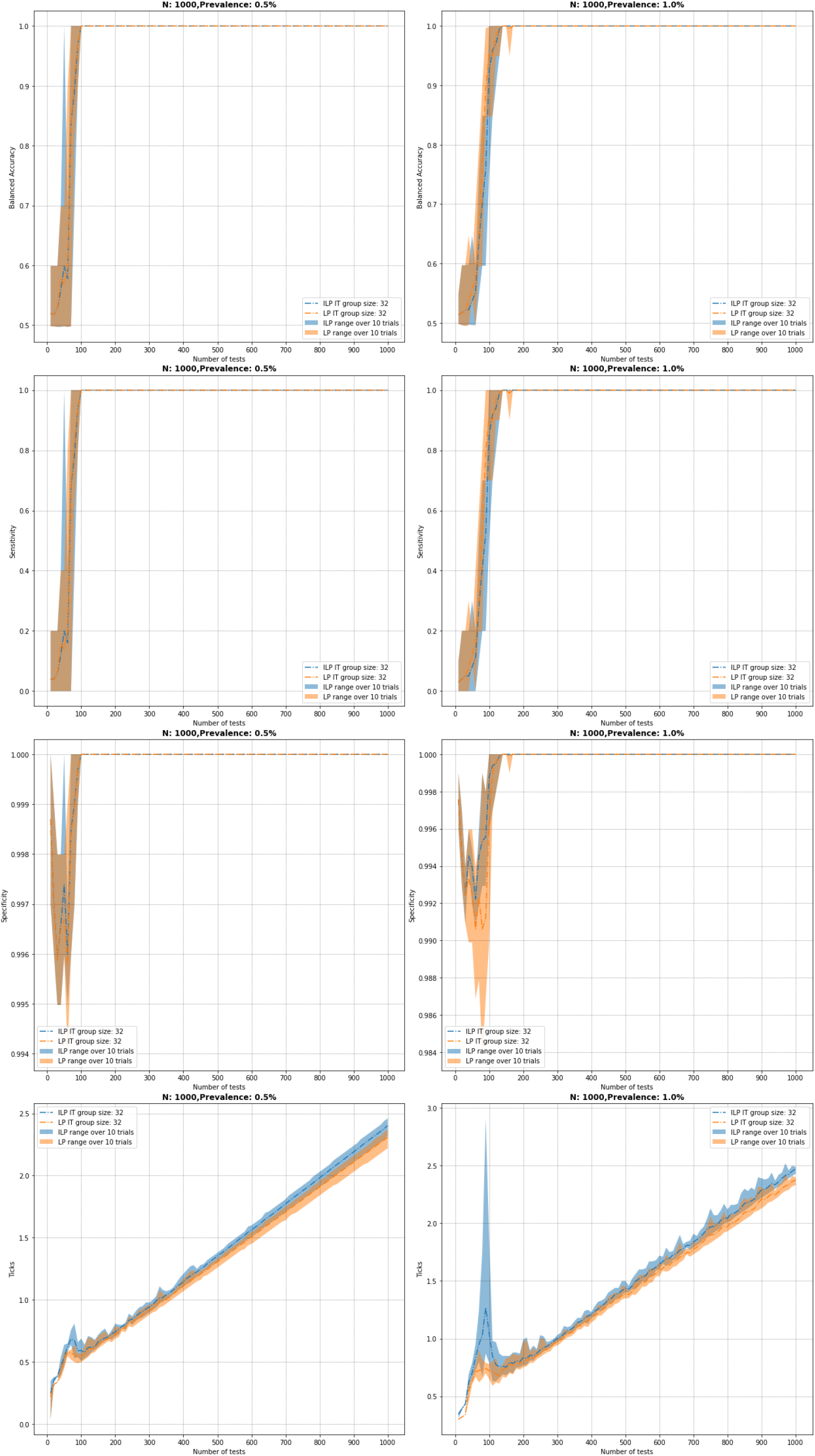
Comparison of (1^st^ **row)** balanced accuracy, (2^nd^ **row)** sensitivity, (3^rd^ **row)** specificity, and (4^th^ **row)** system time (measured in ticks) to fit when *N* = 1000, *g* = 32, and **(left)** *p* = 0.5% and **(right)** *p* = 1%.

Figure 9 gives the same comparison as Figure 8 when *N* = 1000, and *g* = 14, *p* = 5%, and also *g* = 7, *p* = 10%. As in the previous figure, we can see a sharp decrease in specificity of the LP decoder near the point where the balanced accuracy crosses 0.75 (approximately *m* = 200 tests when *p* = 5% and *m* = 350 tests when *p* = 10%). We also note corresponding increases in the system times to solve near these transition points.

**Figure 9:**
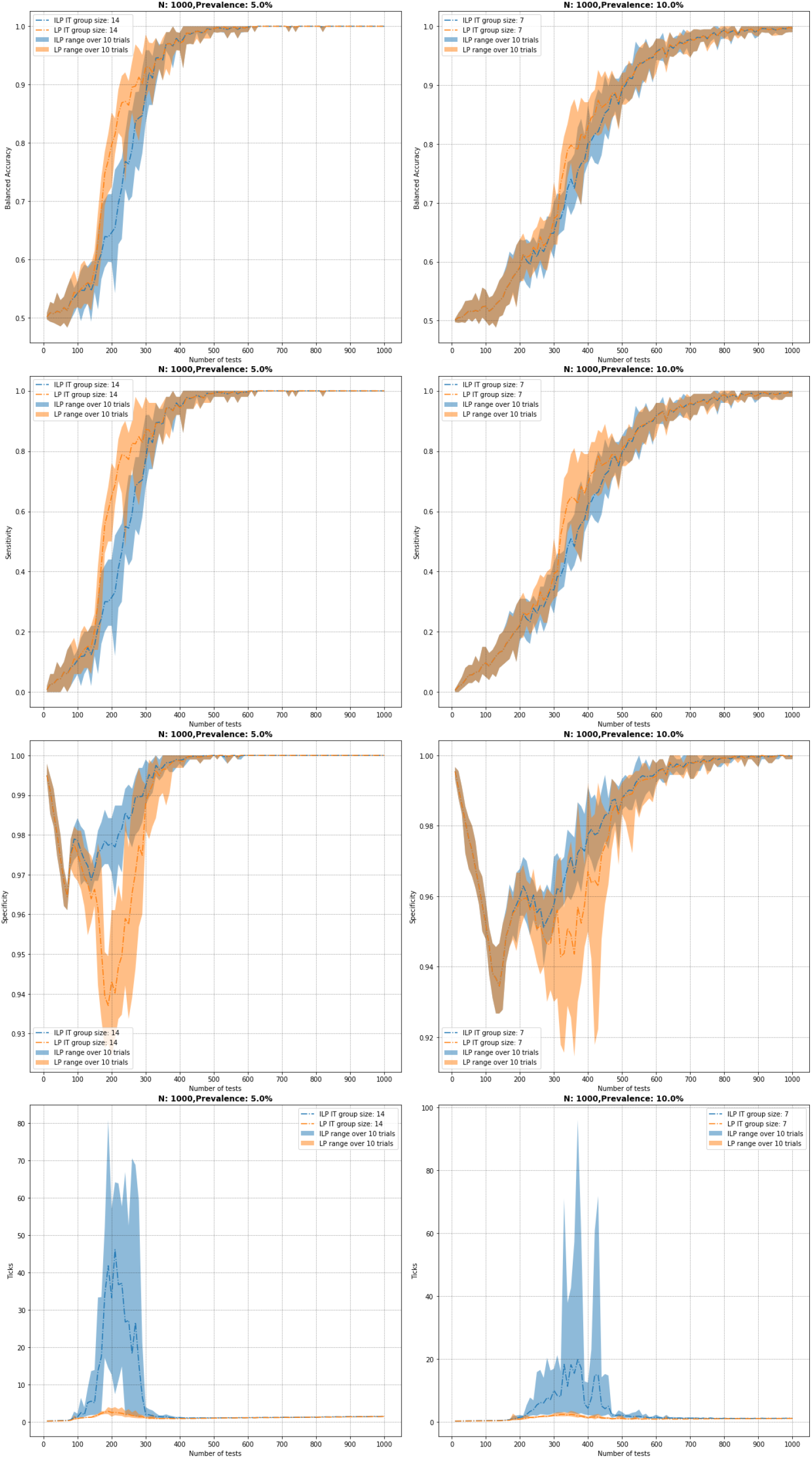
Comparison of (1^st^ **row)** balanced accuracy, (2^nd^ **row)** sensitivity, (3^rd^ **row)** specificity, and (4^th^ **row)** system time (measured in ticks) to fit when *N* = 1000, and **(left)** *g* = 14, *p* = 5 and **(right)** *g* = 7, *p* = 10.

Finally, Figure 10 gives the same comparisons as the last two figures when *N* = 1000, and *g* = 4, *p* = 15%, and also *g* = 3, *p* = 20%. Here we note the sharp decline in specificity of both methods until approximately *m* = 250 in the case *p* = 15% and *m* = 325 in the case *p* = 20% group tests are reached. We also do not observe a significant decrease in specificity of the LP near the previously mentioned transition point when the balanced accuracy exceeds 0.75, in contrast to the previous two figures where such a decrease was observed. However, we do note the sharp increase in system time required by the ILP at this point when *p* = 15% (approximately *m* = 500 tests). We also observed increased system time to solve for the ILP, peaking at about *m* = 700 tests when the balanced accuracy crosses 0.85.

**Figure 10:**
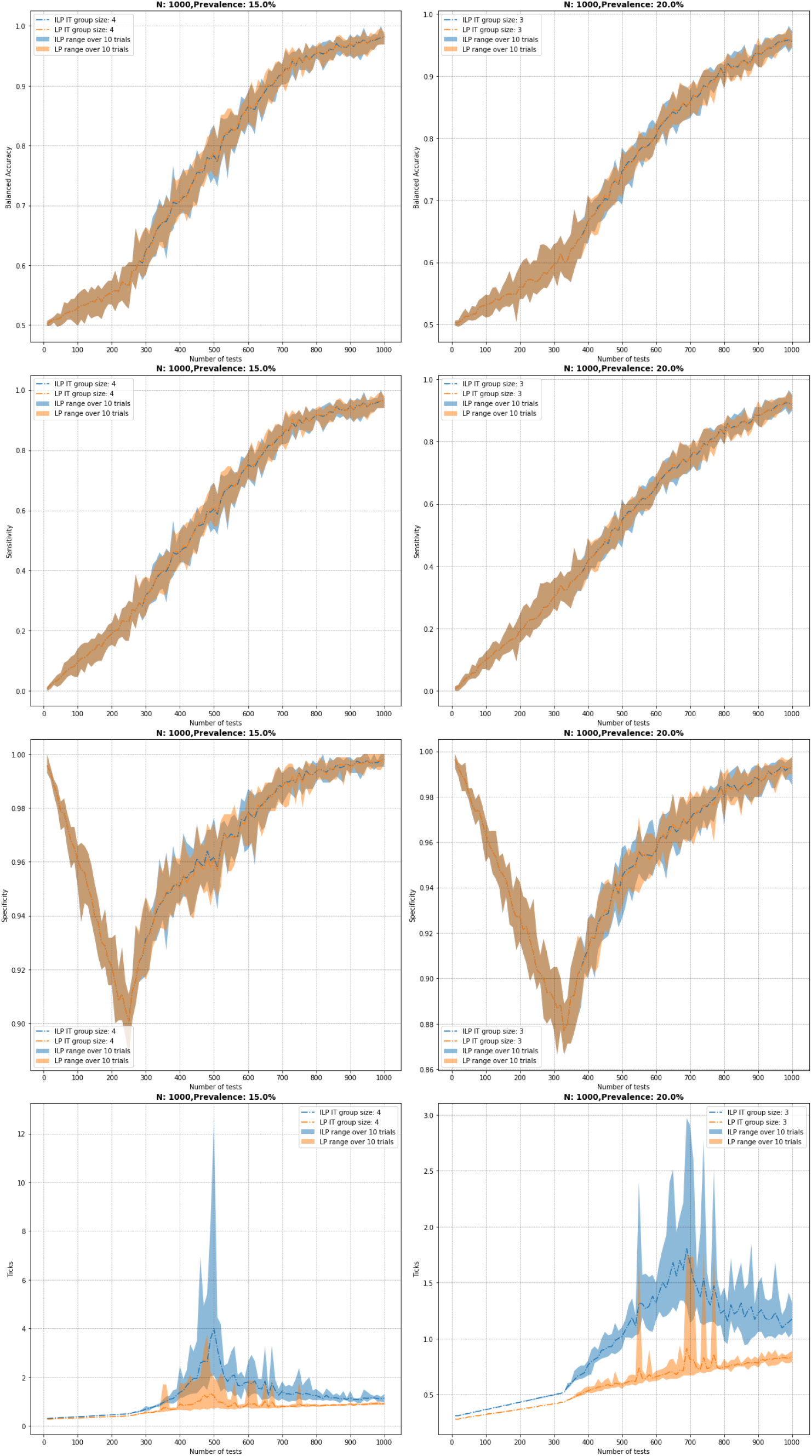
Comparison of (1^st^ **row)** balanced accuracy, (2^nd^ **row)** sensitivity, (3^rd^ **row)** specificity, and (4^th^ **row)** system time (measured in ticks) to fit when *N* = 1000, and **(left)** *g* = 4, *p* = 15% and **(right)** *g* = 3, *p* = 20%.

We summarize our observations on the efficacy of solving the ILP in comparison to solving the LP, with the choice of rounding threshold *β* = 0, as follows. For larger group sizes and smaller prevalences, e.g., *p* = 0.5%, 1%, 5%, or 10% and *g* ≥ 7, the LP approach often provides marginally better balanced accuracy and sensitivity at the expense of worse (and sometimes substantially worse) specificity near the observed transition points. The ILP approach expends additional computational effort near these points to identify true negatives, as can be seen by the increased system time required to solve, hence yielding better specificity. Away from these transition points, both approaches provide similar results in terms of the measured statistics and solve time. For higher prevalences and smaller group sizes, e.g., *p* = 15% or *p* = 20% and *g* = 4 or *g* = 3, the ILP and LP approaches provide similar results in the measured statistics, with the ILP taking marginally longer to arrive at a solution. We note that these results depend heavily on the choice of threshold parameter *β*. The **GroupTesting** package provides an input variable for this parameter, allowing for the rounding threshold to be tuned for the given sensitivity and specificity trade-off requirements in practical applications.

Note that the comparison in the appendix A are for noiseless setting. It also worth mentioning the precise format of the LP relaxation for the ILP (4). This is defined as

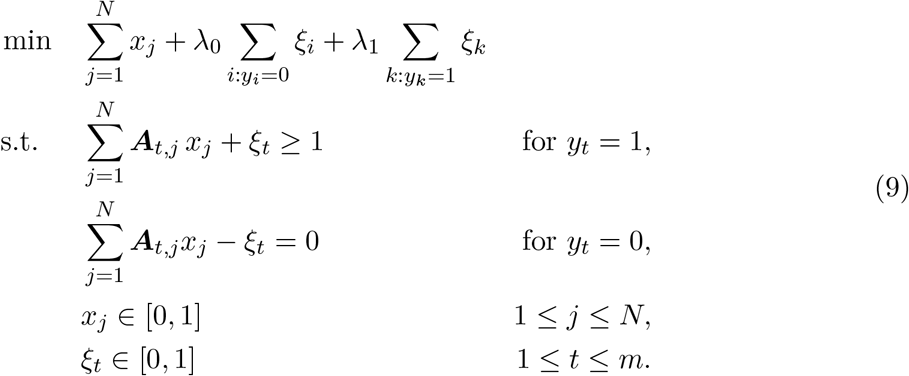

This formulation is also available in the **GroupTesting** package.

### B. Cross-validation for selecting regularization parameters

In this section we briefly discuss the cross-validation procedure employed by the **GroupTesting** package. As discussed in Section 5, **GroupTesting** supports cross-validation for both *λ*_*e*_ (in the case *λ*_0_ = *λ*_1_ = *λ*_*e*_) or for both parameters *λ*_0_ and *λ*_1_ independently, selecting the pair (*λ*_0_, *λ*_1_) yielding the best result. The cross-validation procedure divides the test data into *K* equal parts, selecting one part for training and using the remaining *K −* 1 parts for testing. Here we use *K* = 5 folds for cross-validation. We then cycle through each pairing of testing and training splits, solving the group testing problem with values *λ*_*e*_ ∈ {1*/*8, 1*/*4, 1*/*2, 1, 2, 4, 8}and choosing the values leading to the lowest averaged testing errors, i.e. that predicts the results of the group tests with lowest error, taken here to be the balanced accuracy score. Figure 11 displays the results obtained by this procedure. There we see the cross-validation procedure consistently chooses a value for *λ*_*e*_ providing accuracies near the best-achievable over a range of alternative fixed parameter values.

**Figure 11:**
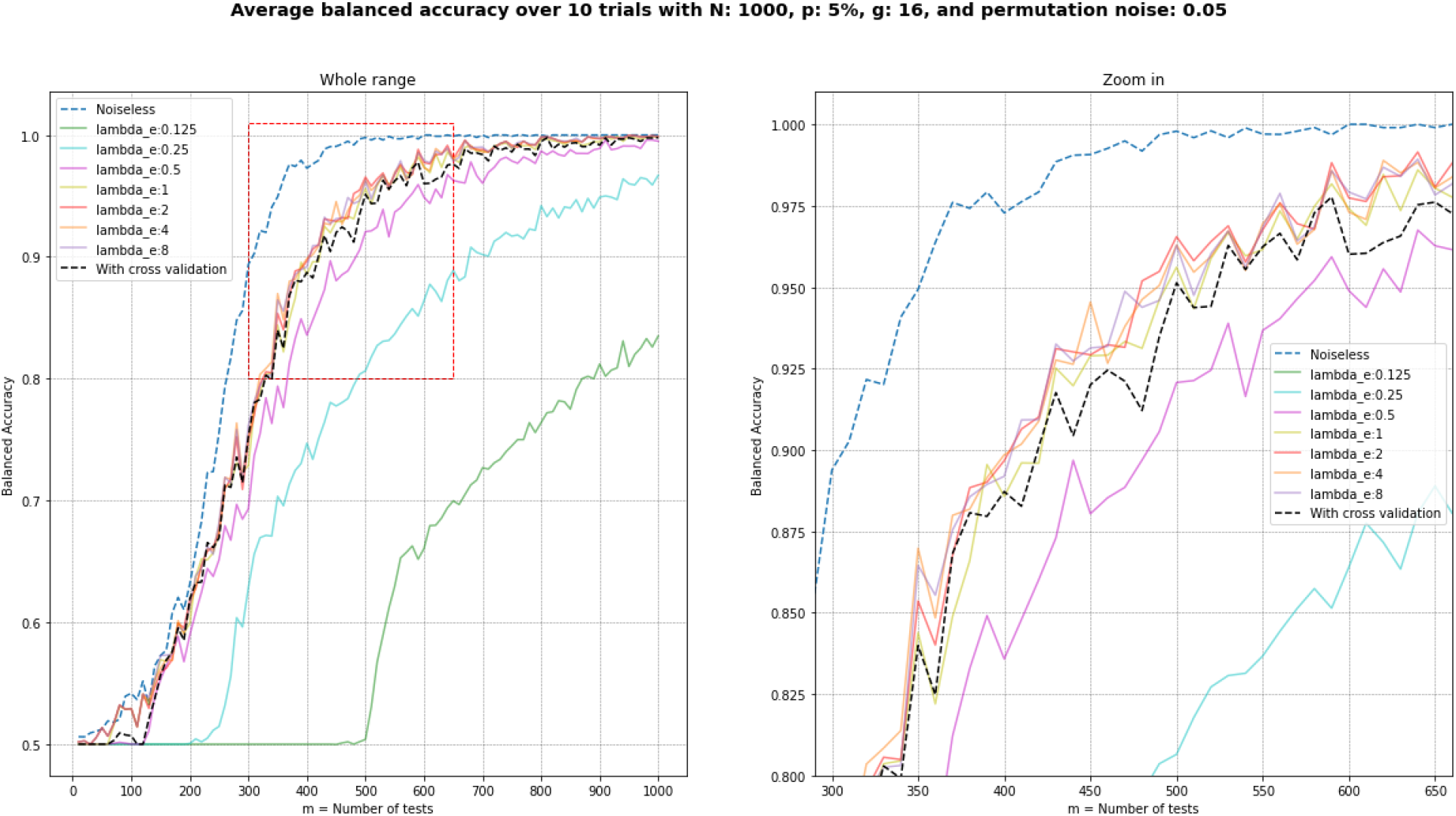
**Left:** Comparison of average balanced accuracy with *N* = 1000, prevalence *p* = 5%, group size *g* = 16, and 5% permutation noise over a range of values of *λ*_*e*_ and compared to those selected with cross-validation, **right:** same figure zoomed in. Results obtained on the noiseless problem are included for comparison.

## Notes

### Competing Interest Statement

The authors have declared no competing interest.

### Author Declarations

All data in this manuscript generated via the pipeline introduced in the manuscript and this manuscript does not contain any human/patient/participant data.

